# Comparison of the Distribution of the Dental Hygienist Workforce and Population in Ontario: A Geospatial Analysis

**DOI:** 10.1101/2025.04.23.25326253

**Authors:** Mark J. Dobrow, Eric Bruce, Keisha Simpson, Glenn Pettifer

## Abstract

**Objectives:** This study conducted a geospatial analysis of the distribution of the dental hygienist workforce relative to the distribution of the population in Ontario, Canada, aiming to address workforce imbalances and inform regional and international workforce planning.

**Methods:** Geospatial analysis techniques were employed to examine the dental hygienist workforce distribution using anonymized datasets from the College of Dental Hygienists of Ontario (the professional regulatory body) and the Canadian census. The data were linked using the forward sortation area (FSA) component of Canadian postal codes, covering 520 FSAs across Ontario. Analyses were conducted at three levels, based on different aggregations of postal code data.

**Results:** The study found significant variations in the distribution of dental hygienists across Ontario. The analysis revealed pockets of high dental hygienist density, mostly in urban areas, and areas with low dental hygienist rates, especially in rural and remote locations. The overall provincial rate was 97 dental hygienists per 100,000 population, with variation across the 520 FSAs, from zero to 20,000 dental hygienists per 100,000 population (or zero to 739 dental hygienists per 100,000 population if five outlier FSAs were removed).

**Conclusions:** The study underscores the complexity of dental hygienist workforce distribution in Ontario, revealing significant geographical disparities that suggest areas of both oversupply and undersupply. These insights provide actionable guidance for workforce policies and regulatory strategies, such as targeted incentives and public health initiatives, to address the mismatch between workforce supply and population needs. The findings highlight the importance of regular geospatial analyses to track changes in workforce distribution over time. The rigorous methodological approach and comprehensive evaluation of potential limitations offer valuable guidance for similar analyses in other jurisdictions. By providing a detailed framework and insights that extend beyond Ontario, this study contributes to the global understanding of dental hygienist workforce dynamics and supports the development of informed policies on a broader scale.

## 1. Background

Over the last five years, health systems have encountered significant disruptions. The COVID-19 pandemic has profoundly affected populations and health systems, extending beyond the direct morbidity and mortality caused by the infectious disease. Economic conditions, including inflationary pressures, and migration patterns influenced by the pandemic and global conflicts have posed challenges for various labour markets^1–3^. The shifting work-home dynamic, with a growing number of non-health sector workers partially or fully working from home, has altered demand for many professional health services, moving from areas near workplaces to those closer to homes^4,5^. Health care workforces have been under immense strain, and the prolonged nature of the pandemic, is revealing extended impacts, including potential structural changes in the size and distribution of many health care professions^6–9^.

Understanding workforce distribution is an important step in evaluating whether health resources are being allocated appropriately, effectively, efficiently, and equitably. These patterns can reveal mismatches between workforce supply and population needs, which can inform strategies to improve access to care and optimize resource use. Insights from such analyses can also contribute to workforce planning efforts beyond the immediate context, as similar challenges are often observed in other regions.

The dental hygiene profession has historically been influenced by various workforce dynamics, including regulatory, educational, economic, and health-related factors^10–12^. The COVID-19 Pandemic has underscored additional factors affecting workforce dynamics, such as heightened health risks and safety concerns (e.g., dental hygienists faced high exposure risks due to their work, leading to increased safety concerns and necessary adjustments in the work environment), economic impacts (e.g., the pandemic affected spending on oral health and led to temporary closures or reduced operations in dental practices), and the profession’s adaptability and resilience (e.g., rapid adoption of enhanced safety protocols)^13–15^. For example, research indicates that the United States (US) may be experiencing a shortage of dental hygienists post-pandemic, attributed to factors such as workforce attrition, changes in work-life preferences, and ongoing health and safety concerns^16^. However, it remains uncertain whether these impacts are unique to specific jurisdictions or more widespread globally.

Given these dynamics, there is a pressing need for more detailed information on the current distribution of the dental hygienist workforce and how it aligns with population need. The Canadian Institute for Health Information (CIHI) conducts regular health workforce analyses for Canada, and its most recent data indicated that Ontario had 13,560 dental hygienists in 2021, or nearly 92 dental hygienists per 100,000 population, the highest of any province or territory in Canada^17^. However, CIHI’s analyses do not delve into the intraprovincial distribution of the dental hygienist workforce or compare it to the general population distribution, which serves as a proxy for the demand for dental hygiene services. The absence of detailed demand/supply data complicates the task of regulatory bodies in fine-tuning their approaches to registering dental hygienists. Consequently, this study aimed to perform a geospatial analysis of the distribution of the dental hygienist workforce relative to the population distribution in Ontario, Canada.

## 2. Methods

Geospatial analysis refers to the process of examining and interpreting data that has a geographic component^18^. This involves using tools and techniques to collect, store, analyze, and visualize data that is tied to a specific geographic location, such as global positioning system (GPS) coordinates, a street address, or postal codes. The goal of geospatial analysis is to gain a better understanding of the spatial relationships and patterns within the data, and to use this information to take more informed actions.

Geospatial analyses of oral health care workforces are not common. Studies we identified focus on a mix of high– and middle-income countries (e.g., US^19^, Australia^20^, Canada^21^, Germany^22^, Kenya^23^, Malaysia^24^ and Sri Lanka^25^), with each assessing geographic access to oral health professions. A recent narrative review^26^ explored the utilization of geographic information systems (GIS) in dental public health research, describing a useful framework with four domains of GIS studies: (1) measurement (“gauging the distribution of communicable and non-communicable diseases in an area and the environment’s effect on them”), (2) mapping (“developing maps that portray characteristics and help in the spatial understanding of a population’s health”), (3) monitoring (“monitor changes in health and diseases in space and time”), and (4) modelling (“to model alternatives of actions and process operations based on the risk prediction of diseases”)^26^. Each of the studies noted above are consistent with the mapping domain, which has been further characterized as “[a]lso gaug[ing] the healthcare access and spatial distribution of healthcare providers”^26^. While this aligns closely with our aims, the review also includes a valuable cautionary note that “[a]ccess to health care is different from the geographic accessibility, in that the former encompasses both spatial components (availability and accessibility) and aspatial components (acceptability and affordability). The latter, that is, the geographic accessibility, comprises only the spatial components”^26^. Thus, the approach to geospatial analysis taken for this study focuses specifically on geographic accessibility of the dental hygienist workforce in Ontario. The data sources, data linkage and levels of analysis are outlined below.

### 2.1 Data Sources

This project used two main data sources that each contained geographic information, including (1) an anonymized dataset from the College of Dental Hygienists of Ontario (CDHO) that contains administrative and demographic data on its dental hygienist registrants, including postal codes, and (2) the most recent publicly available Canadian census data from Statistics Canada, which has a version that includes postal code information. The CDHO dataset is essentially a cross-section of the registrant database taken at a specific point in time, in this case on 02 May 2023, that includes practice and/or home postal codes plus a range of variables including membership status, sex, age, graduation date, university/college country/province and education level attained. As the CDHO dataset captured multiple practice locations for some dental hygienists, their primary practice location postal code was used, and when unavailable, their home postal code. Similarly, the Statistics Canada census dataset provides a cross-section of a limited set of population variables at another period in time, in this case for the 2021 calendar year (the 2021 census was released on 09 February 2022)^27^. Statistics Canada also generates a boundary file that contains the necessary geographic coordinates to map the postal code data included within the census dataset^28^.

### 2.2 Data Linkage: Forward Sortation Area (FSA)

To link the CDHO dataset to the census dataset, the common postal code data field included in each was matched. More specifically, the ‘forward sortation area’ (FSA) component of the full postal code, which was available in both datasets, was used. The FSA is composed of the first three characters of all six-character Canadian postal codes (that alternate alpha-numeric characters). For example, the ‘M1A’ FSA includes all available postal codes under that prefix (i.e., from M1A 0A0 to M1A 9Z9). While Canada Post is constantly updating the FSAs, as of their update in February 2023^29^, there were 532 FSAs in Ontario (see **Additional file 1** for a complete list of Ontario FSAs and their Canada Post location descriptors).

It is important to note that while the FSA provides a means to link otherwise disparate datasets and facilitate geospatial analysis, it was not established for this purpose. Rather, the FSA was established by Canada Post to optimize mail sorting and delivery. While the FSA represents a valuable option for conducting geospatial analyses, two key limitations should be considered. First, FSA boundaries are defined based on postal routes rather than natural, administrative, or socio-political boundaries, which can create some non-intuitive geographic regions that may cut across municipalities/neighbourhoods or may include one or more non-contiguous areas. Second, FSAs can vary widely in terms of geographic and population size. Some FSAs cover large rural or remote regions with sparse populations (which Canada Post usually designates with a zero for the second character of the FSA), while other FSAs represent very small but densely populated urban areas. Despite these limitations, FSAs are a widely used and valuable tool for geospatial analysis, and understanding these limitations informs how resultant analyses are interpreted and used.

One final note on FSAs. Although there are currently 532 FSAs in Ontario, we were not able to include all of them in our final dataset. The Statistics Canada census dataset contained population data for 521 Ontario-based FSAs (the remaining 11 FSAs had zero population counts which do not allow a dental hygienist rate to be calculated), while Statistics Canada lacked a boundary file for another FSA, leaving a final dataset of 520 FSAs for the analysis. **Additional file 2** identifies and provides the rationale for the 12 FSAs excluded from the analysis.

### 2.3 Three Levels of Analysis

Three levels of analysis were conducted. The main level of analysis focused on the 520 FSAs, however, for some analyses 520 data points makes interpretation more challenging, particularly when examining map outputs.

Therefore, the FSA data were also aggregated to larger regions to maintain the geospatial information and provide additional insights. The second level of analysis, which is referred to throughout this paper as ‘FSA-2’, draws on the first two characters of the FSA (e.g., ‘M1’). This aggregates data from all available FSAs under that prefix (i.e., M1 includes all FSAs from M1A to M1Z). Similarly, the third level of analysis, which is referred to as ‘FSA-1’, draws on the first character of the FSA (e.g., ‘M’). This includes all available FSAs under that prefix (i.e., M includes all FSAs from M0A to M9Z). **Table 1** summarizes the levels of analyses used in this project.

**Table 1:** Levels of Analysis.

| Level of Analysis | FSA | FSA-2 | FSA-1 |
| --- | --- | --- | --- |
| Number of Geographic Areas | 520 FSAs | 50 FSA-2s | 5 FSA-1s |
| Eligible FSAs/FSA-2s/FSA-1s | K0A to K9Z<br>L0A to L9Z<br>M0A to M9Z<br>N0A to N9Z<br>P0A to P9Z | K0-K9<br>L0-L9<br>M0-M9<br>N0-N9<br>P0-P9 | K (Eastern Ontario)<br>L (Central Ontario)<br>M (Metropolitan Toronto)<br>N (Southwestern Ontario)<br>P (Northern Ontario) |

FSA – Forward Sortation Area

## 3. Results

The results are presented in three ways. First, independent summaries of the two main data sources are presented to explore population distribution by FSA, FSA-2, and FSA-1 and dental hygienist distribution by FSA,

FSA-2, and FSA-1. Second, a summary of the analyses of the linked datasets on the distribution of the dental hygienist workforce in Ontario are presented. Third, a series of heat maps to summarize the geospatial analysis are presented.

### 3.1 Independent Summaries of Population and Dental Hygienist Registrant Data Sources

To provide context, independent summaries of the results from the two main datasets are presented: the Statistics Canada census dataset and the CDHO registrant dataset.

#### 3.1.1 Statistics Canada Census Dataset

The Statistics Canada 2021 census dataset indicates that Canada’s population was 36,991,981 in 2021, while Ontario’s population was 14,223,942. As noted earlier in the paper, of Ontario’s 532 FSAs, 11 had a population count of zero and were excluded from the census dataset, while one additional FSA was excluded as Statistics Canada did not provide an associated boundary file to support geospatial analysis. Thus, the population data relates to 520 FSAs. Wide variation by FSA was observed, with the census dataset indicating that the smallest population among the 520 FSAs was only five persons, while the largest population among the 520 FSAs was 115,850. The median and mean population size per FSA was 23,399 and 27,354 respectively, indicating some skewing of the population data towards a few large population FSAs.

**Additional file 3** presents the population data by FSA (**Figure AF3a**), FSA-2 (**Figure AF3b**) and FSA-1 (**Figure AF3c**), each ordered by smallest to largest population. The figures highlight the overall variation in population size by FSA and **Figure AF3b** and **Figure AF3c** emphasize the population differences across FSA-2 and FSA-1 areas, with ‘L’ FSAs (central Ontario) containing a greater relative proportion of the Ontario population and ‘P’ FSAs (northern Ontario) containing a smaller relative proportion of the Ontario population. **Additional files 4-6** provide the specific population count for each FSA, FSA-2, and FSA-1.

#### 3.1.2 CDHO Registrant Dataset

The CDHO registrant dataset included 22,587 records. To be eligible for the analyses, each record needed to have (1) a membership status of ‘general’ or ‘specialty’, which indicated active/practicing dental hygienists, and (2) an Ontario-based postal code. It was not possible to determine accurately from the CDHO dataset what portion of these dental hygienists worked full-time. Ultimately, 13,856 records met the eligibility criteria and were included in the analyses (the majority of the remaining 8,731 dental hygienist members’ status was resigned, inactive, revoked, deceased, or suspended).

**Table 2** provides descriptive results for the 13,856 active/practicing dental hygienist registrants. The vast majority are female (97%), holding a ‘general’ membership status (96%), and received their university/college education in Canada (96%). The basic education levels are evenly split with 52% holding a degree, and 48% holding a diploma. Age and years since graduation are skewed towards dental hygienists 40 years of age and younger (53%) and dental hygienists who graduated within the last 20 years (74%) (**Figure 1**).

**Figure 1:**
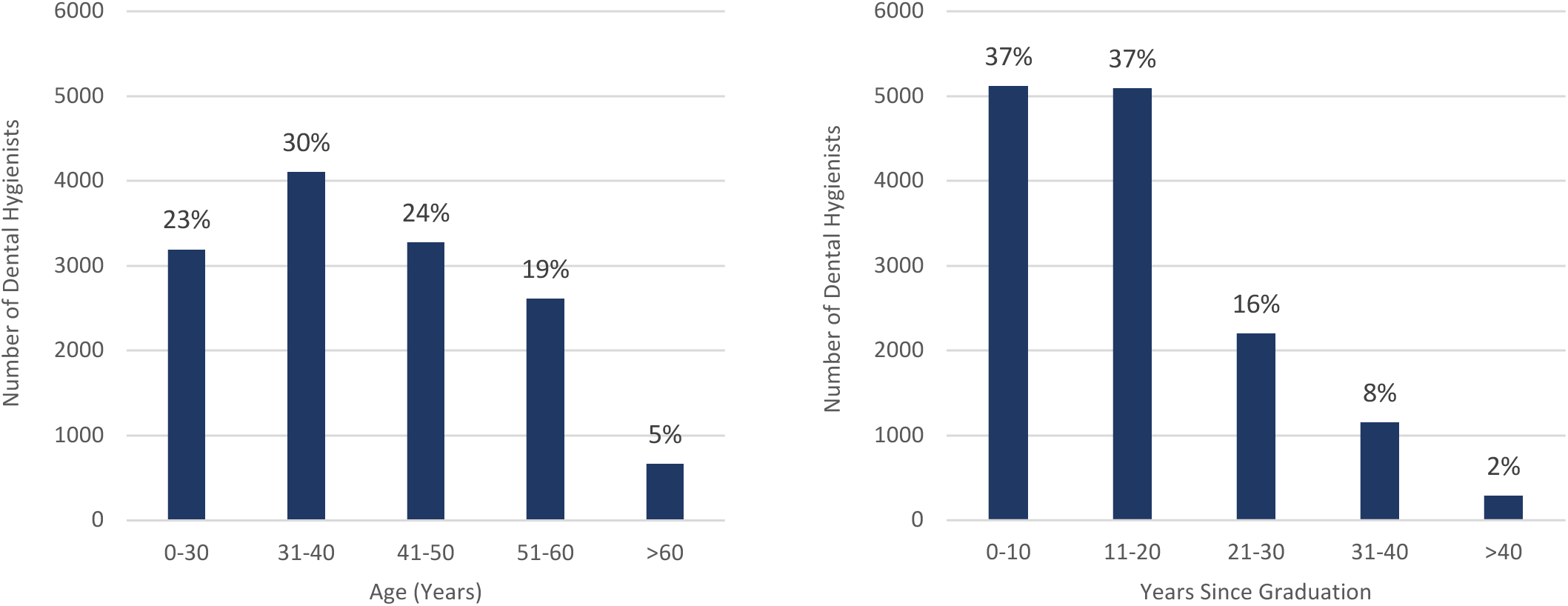
Ontario Dental Hygienist by Age and Years Since Graduation.

**Table 2:** CDHO Registrant Dataset – Descriptive Results.

| Member Characteristics |  | Characteristic Distribution |  |  |  | Total |
| --- | --- | --- | --- | --- | --- | --- |
| Membership Status | **General:**<br>13,240 (96%) | **Specialty:**<br>616 (4%) |  |  |  | **13,856 (100%)** |
| Sex | **Female:**<br>13,452 (97%) | **Male:**<br>404 (3%) |  |  |  | **13,856 (100%)** |
| Age | **0-30 years:**<br>3,194 (23%) | **31-40 years:**<br>4,106 (30%) | **41-50 years:**<br>3,277 (24%) | **51-60 years:**<br>2,612 (19%) | **>60 years:**<br>667 (5%) | **13,856 (100%)** |
| Years Since Graduation | **0-10 years:**<br>5,120 (37%) | **11-20 years:**<br>5,094 (37%) | **21-30 years:**<br>2,202 (16%) | **31-40 years:**<br>1,153 (8%) | **>40 years:**<br>287 (2%) | **13,856 (100%)** |
| University/College Country | **Canada:**<br>13,304 (96%) | **USA:**<br>385 (3%) | **UK:**<br>47 (0.3%) | **Other:**<br>120 (1%) |  | **13,856 (100%)** |
| Basic Education Level | **Degree:**<br>7,245 (52%) | **Diploma:**<br>6,611 (48%) |  |  |  | **13,856 (100%)** |

CDHO – College of Dental Hygienists

Similar to the population data, when the dental hygienist data were assessed by FSA, wide variations were observed, with 27 FSAs having no dental hygienists, while the most dental hygienists per FSA was 137. The median and mean number of dental hygienists per FSA was 22 and 27 respectively, indicating some skewing of the dental hygienist data towards a few FSAs with higher numbers of dental hygienists. **Additional file 7** presents the CDHO registrant data by FSA (**Figure AF7a**), FSA-2 (**Figure AF7b**) and FSA-1 (**Figure AF7c**), each ordered by the smallest to largest number of dental hygienists. The figures highlight the overall variation in the number of dental hygienists by FSA and **Figure 3b** and **Figure 3c** emphasize the differences across FSA-2 and FSA-1 areas, with ‘L’ FSAs (central Ontario) containing a greater relative proportion of Ontario dental hygienists and ‘P’ FSAs (northern Ontario) containing a smaller relative proportion of the dental hygienists in the province. **Additional files 4-6** provide the specific dental hygienist counts for each FSA, FSA-2, and FSA-1.

### 3.2 Summary Results for the Distribution of Dental Hygienist Workforce Based on Linked Datasets

Although there were 13,856 dental hygienists in the CDHO database that met the eligibility criteria for the project, 13 were from four FSAs (L3J, M5K, M5X, M7A) that were excluded from the linked dataset, as Statistics Canada had no population data for three (L3J, M5K, M5X) nor boundary coordinates for the fourth (M7A). L3J is a new FSA that Canada Post established in 2022, therefore was not included in the 2021 census dataset. M5K and M5X are both one block areas in a business zone of downtown Toronto containing office towers. M7A is the FSA for the Ontario legislature and accompanying office buildings known as ‘Queen’s Park’ in Toronto.

Therefore, for the remainder of the analyses presented in this report, the focus is on 13,843 dental hygienists distributed across 520 FSAs. The first way that the linked dataset was assessed was to compare the distribution of the population across FSAs to the distribution of dental hygienists across FSAs in the province. **Figure 2** overlays the population data and dental hygienist data on the same chart for FSAs (**Figure 2a**), FSA-2s (**Figure 2b**), and FSA-1s (**Figure 2c**). If there was a perfect relationship between population size and the number of dental hygienists, you would see the navy bars and orange bars align with one another. However, as **Figure 2a** shows, when assessing the 520 FSAs, there is considerable variation, with FSAs with similar populations having very different numbers of dental hygienists. As the data are aggregated to the FSA-2 and FSA-1 levels, **Figure 2b** and **Figure 2c** show that the variation is masked, and the number of dental hygienists tracks more closely with the corresponding FSA-2 or FSA-1 populations. These results suggest that at more local levels, based on the 520 distinct regions across the province, the number of dental hygienists is variable and inconsistent. There may be reasonable explanations for some of this variability, for example, some of the smaller population FSAs may be situated either in small urban areas (e.g., commercial office towers) where no one lives but many dental hygienists work, or in large rural or remote areas that have very few dental hygienists. However, to understand the underlying factors that impact on the inconsistencies in the number of dental hygienists across Ontario, further investigation is required.

**Figure 2a:**
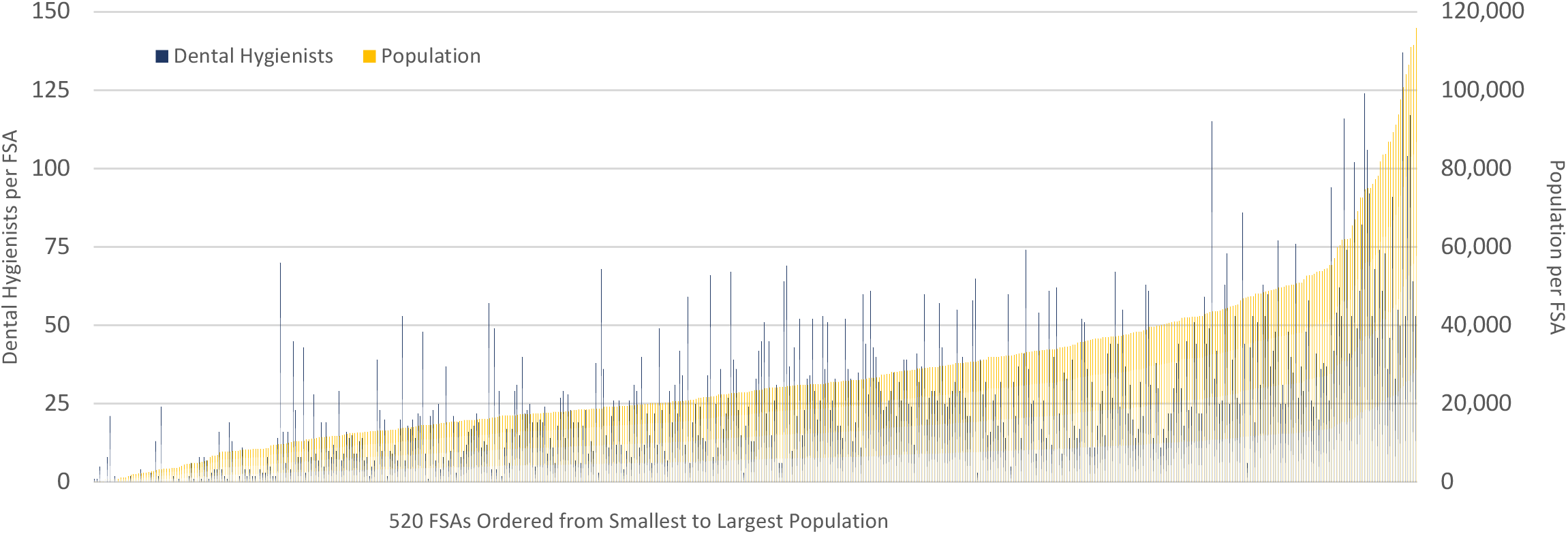
Dental Hygienists and Population per Forward Sortation Area (FSA) in Ontario.

**Figure 2b:**
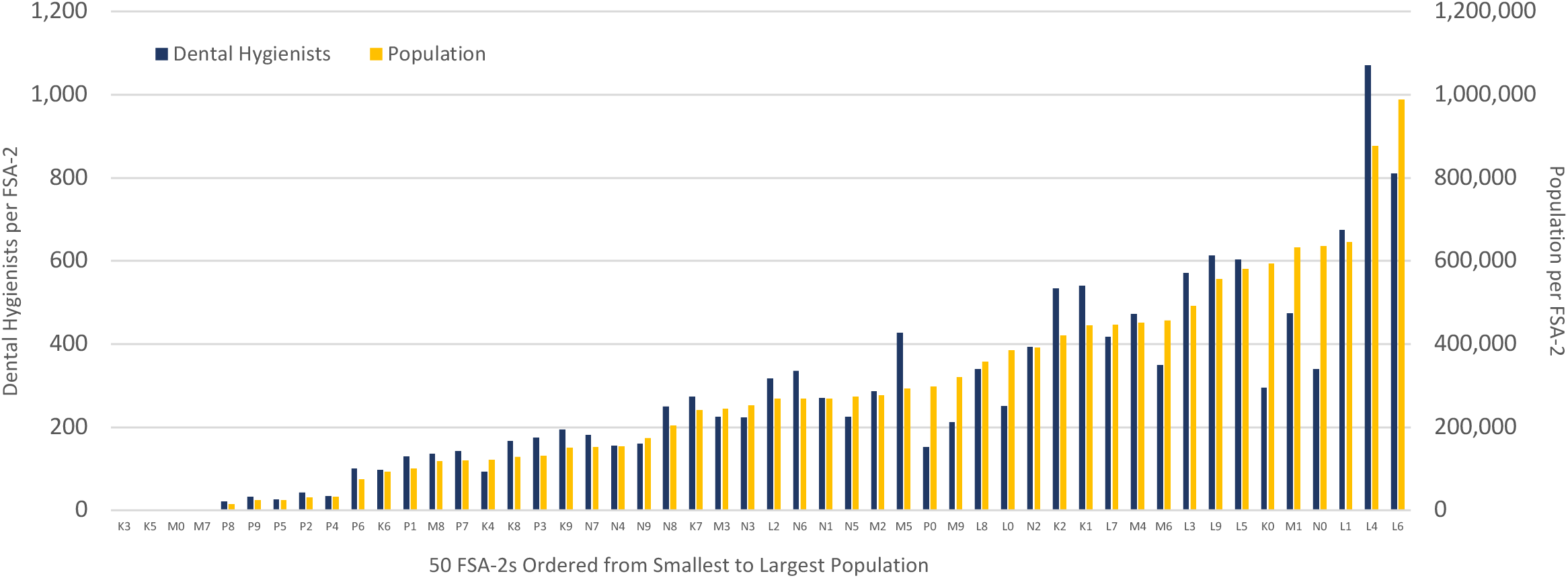
Dental Hygienists and Population per Forward Sortation Area-2 (FSA-2) in Ontario.

**Figure 2c:**
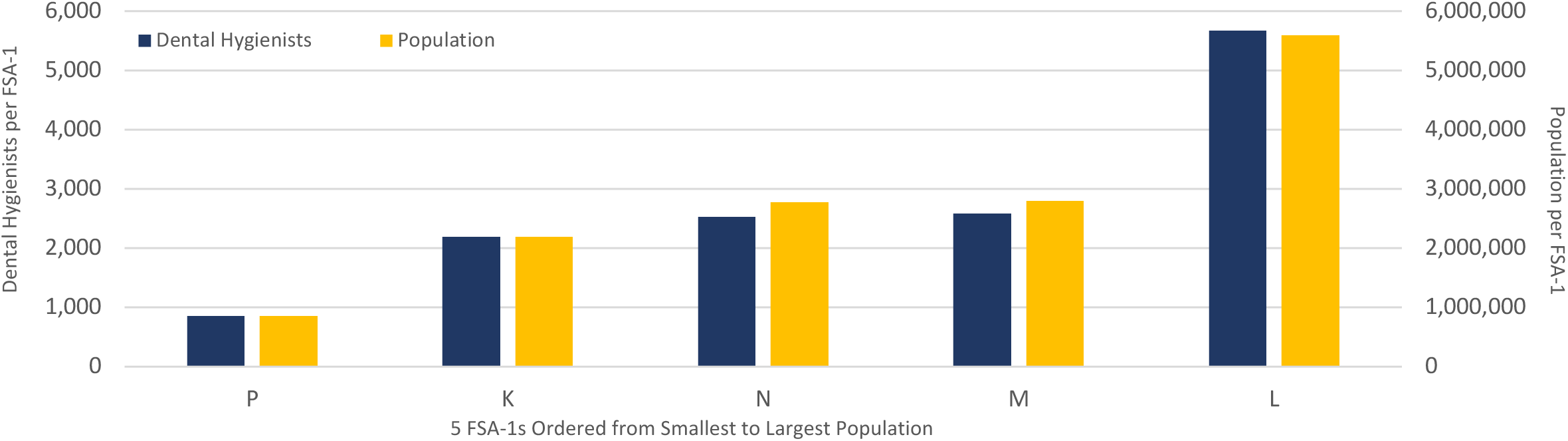
Dental Hygienists and Population per Forward Sortation Area-1 (FSA-1) in Ontario.

The second way that the linked dataset was assessed was to calculate a dental hygienist rate (i.e., the number of dental hygienists per 100,000 population) per FSA. As with the data on the population and number of dental hygienists for each FSA, the dental hygienist rate also varied widely by geographic region. **Figure 3** presents the dental hygienist rate for FSAs (**Figure 3a1**), FSA-2s (**Figure 3b**), and FSA-1s (**Figure 3c**). Again, if there was a perfect relationship between population size and the number of dental hygienists, you would expect the dental hygienist rate to be consistent for all FSAs, FSA-2s, or FSA-1s. **Figure 3a1** suggests this is not the case, with a few outliers contributing to a broad range of dental hygienist rates. Twenty-seven FSAs had a dental hygienist rate of zero, while the highest dental hygienist rate was 20,000 dental hygienists per 100,000 population. It is clear that these data are impacted by a few outlier results. Only five FSAs have a dental hygienist rate over 750 dental hygienists per 100,000 population, with three of these five FSA’s having total populations under 30 persons. **Figure 32a** presents the dental hygienist rate with a truncated Y-axis, which removes the effects of the outlier values to allow visualization of the dental hygienist rate across the remaining FSAs. When the results are aggregated to the FSA-2 and FSA-1 levels, we see more consistent dental hygienist rates. For the FSA-2s, the dental hygienist rate ranges from 50 to 146 dental hygienists 100,000 population, while for the FSA-1s, the dental hygienist rate ranges from 91 to 101 dental hygienist per 100,000 population.

**Figure 3a1:**
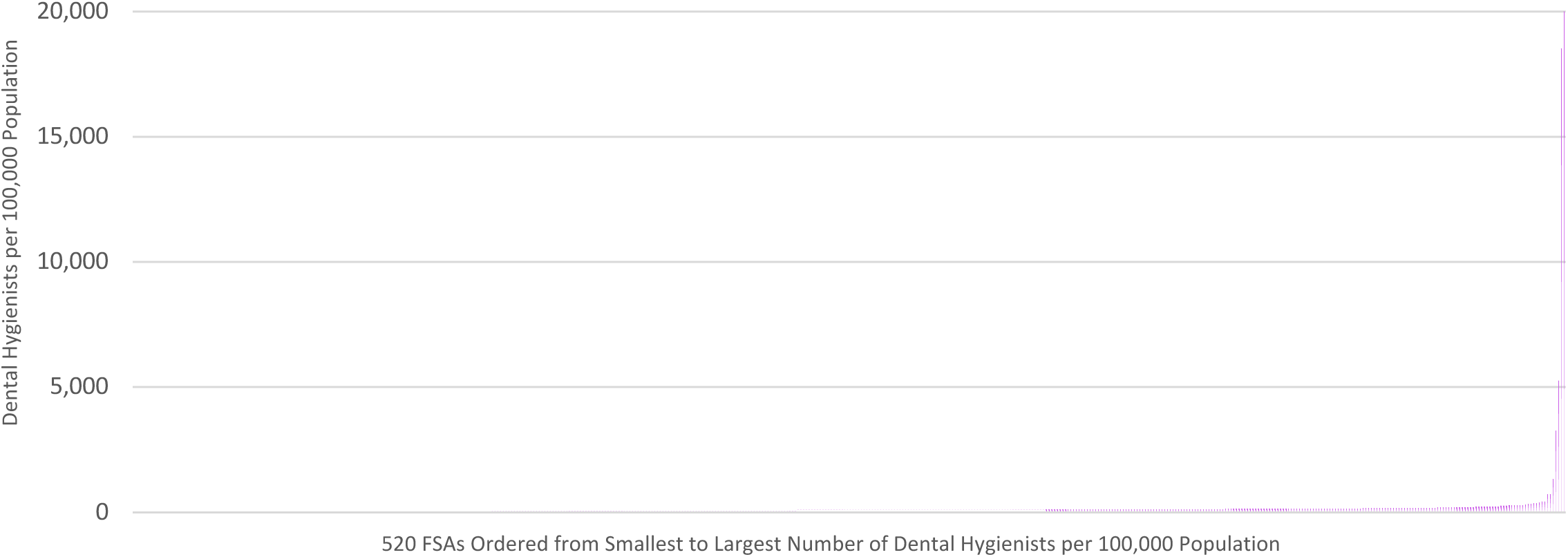
Dental Hygienists per 100,000 Population by Forward Sortation Area (FSA)

**Figure 3a2:**
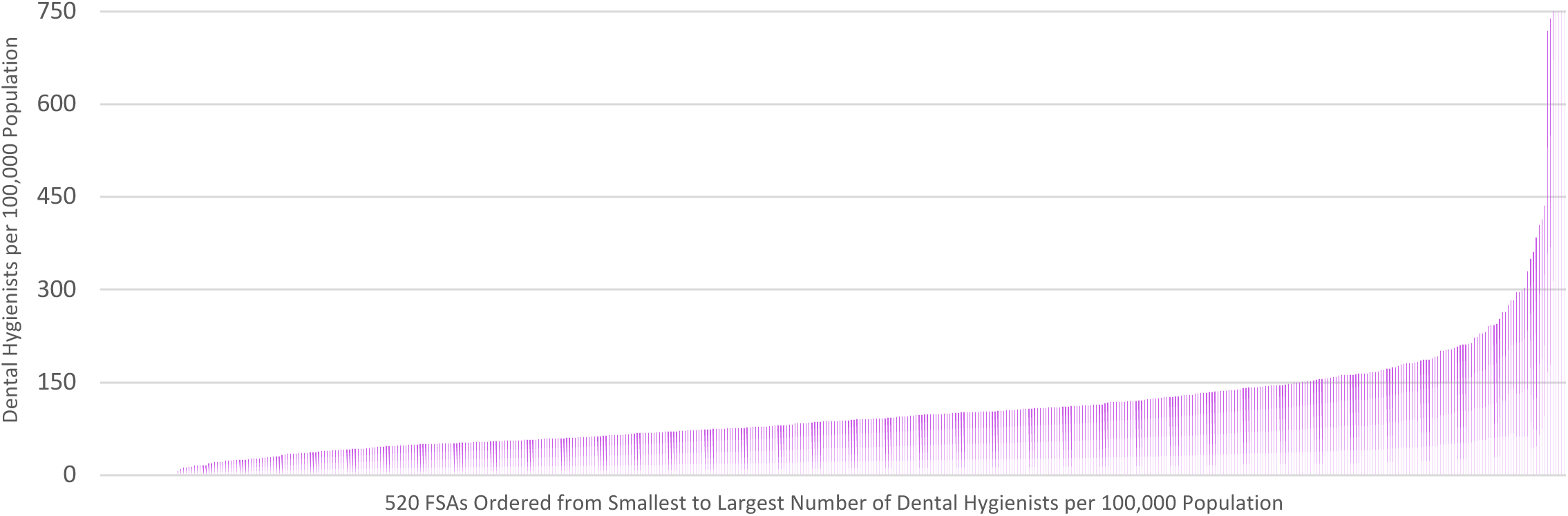
Dental Hygienists per 100,000 Population by Forward Sortation Area (FSA) (Truncated Y-Axis)

**Figure 3b:**
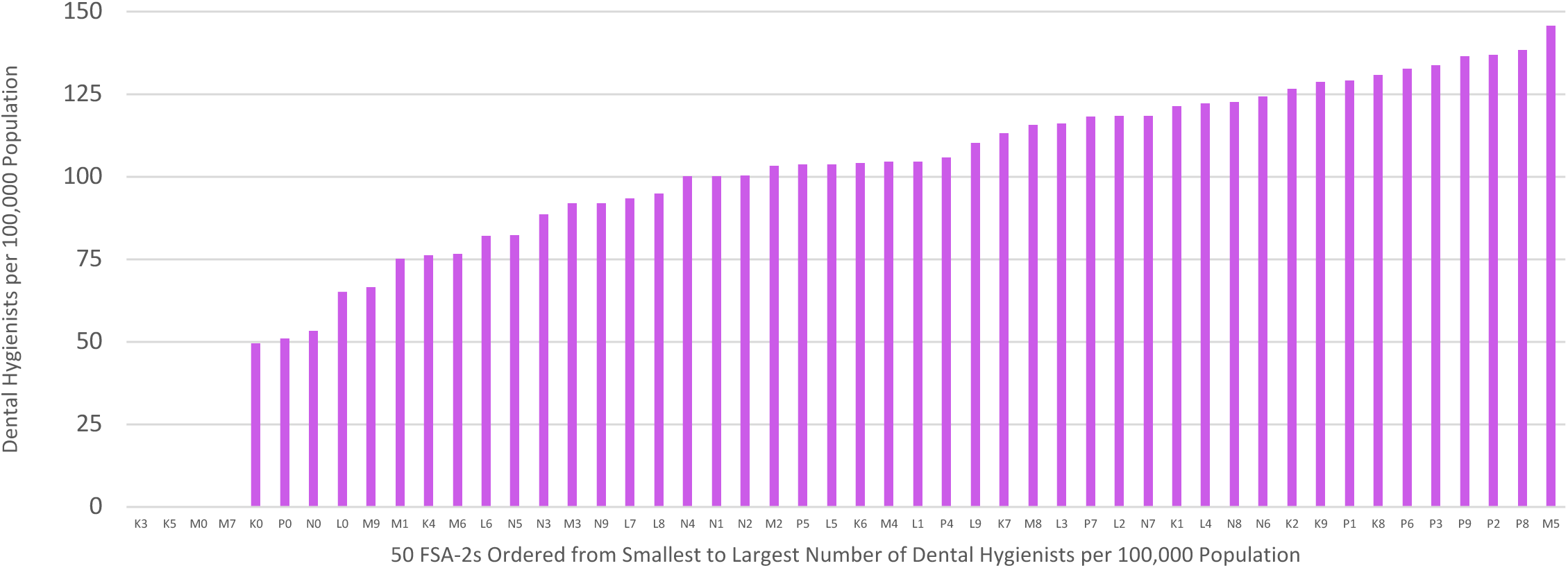
Dental Hygienists per 100,000 Population by Forward Sortation Area-2 (FSA-2)

**Figure 3c:**
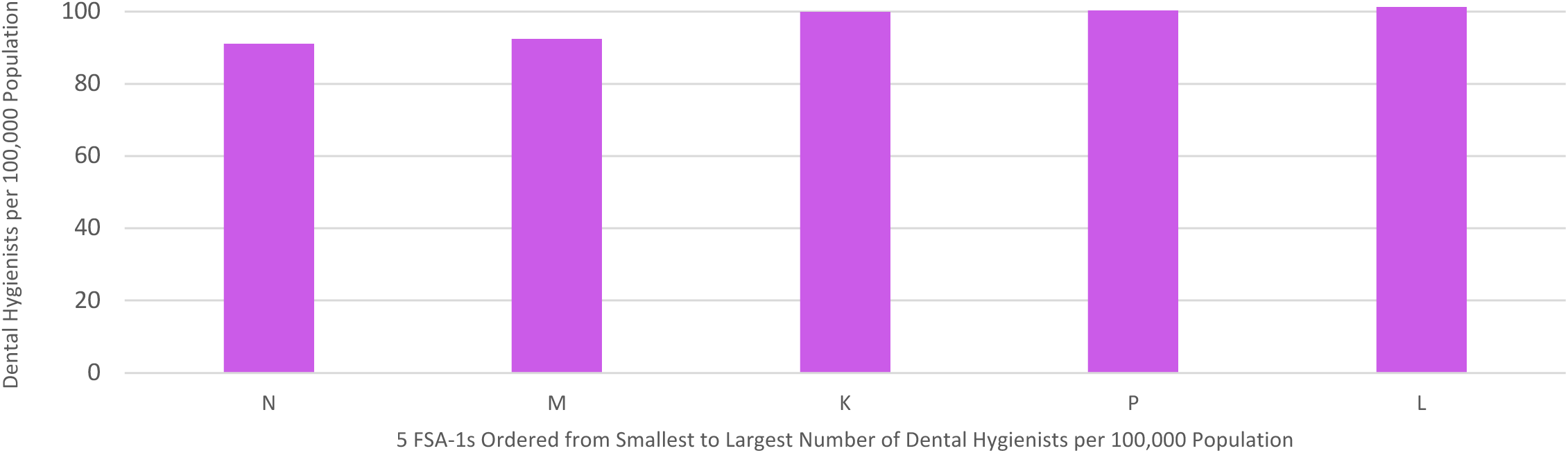
Dental Hygienists per 100,000 Population by Forward Sortation Area-1 (FSA-1)

**Table 3** provides descriptive statistics (measures of central tendency and dispersion) for the three key variables, including the number of dental hygienists, the population, and the dental hygienist rate. **Appendices S4-S6** provide the specific number of dental hygienists per 100,000 population for each FSA, FSA-2, and FSA-1.

**Table 3:** Descriptive Statistics for Number of Dental Hygienists, Population and Dental Hygienist Rate.

|  | Descriptive Statistics | Number of Dental Hygienists per FSA | Population per FSA | Dental Hygienists per 100,000 Population |
| --- | --- | --- | --- | --- |
| **K FSAs**<br>(n=84) | Minimum | 0 | 490 | 0 |
|  | Median | 24 | 20,547 | 98 |
|  | Maximum | 76 | 115,850 | 3256 |
|  | Mean | 26 | 26,154 | 151 |
|  | Standard Deviation | 19.9 | 21,588 | 375.2 |
|  | **Subtotal** | **2,195** | **2,196,966** | **100** |
| **L FSAs**<br>(n=164) | Min | 0 | 5 | 0 |
|  | Median | 28 | 29,215 | 96 |
|  | Max | 137 | 110,956 | 20,000 |
|  | Mean | 35 | 34,142 | 367 |
|  | Standard Deviation | 27.4 | 23,268 | 2,146.2 |
|  | **Subtotal** | **5,671** | **5,599,314** | **101** |
| **M FSAs**<br>(n=96) | Min | 3 | 3,149 | 10 |
|  | Median | 24 | 26,128 | 78 |
|  | Max | 106 | 75,100 | 739 |
|  | Mean | 27 | 29,107 | 117 |
|  | Standard Deviation | 17.3 | 14,002 | 119.3 |
|  | **Subtotal** | **2,586** | **2,794,318** | **93** |
| **N FSAs**<br>(n=118) | Min | 0 | 728 | 0 |
|  | Median | 19 | 20,806 | 82 |
|  | Max | 63 | 86,935 | 384 |
|  | Mean | 21 | 23,555 | 92 |
|  | Standard Deviation | 15.8 | 15,403 | 68.0 |
|  | **Subtotal** | **2,534** | **2,779,475** | **91** |
| **P FSAs**<br>(n=58) | Min | 0 | 96 | 0 |
|  | Median | 8 | 12,216 | 58 |
|  | Max | 74 | 49,727 | 295 |
|  | Mean | 15 | 14,722 | 83 |
|  | Standard Deviation | 18.1 | 9,940 | 70.3 |
|  | **Subtotal** | **857** | **853,863** | **100** |
| **All FSAs**<br>(n=520) | Min |  | 5 | 0 |
|  | Median | 2 | 23,399 | 87 |
|  | Max | 137 | 115,850 | 20,000 |
|  | Mean | 27 | 27,354 | 192 |
|  | Standard Deviation | 22.1 | 19,526 | 1,219.8 |
|  | **TOTAL** | **13,843** | **14,223,936** | **97** |

FSA – Forward Sortation Area

### 3.3 Map Outputs for the Distribution of Dental Hygienist Workforce

To provide a high-level overview of geographic variation in the distribution of dental hygienists across Ontario, two series of standard heat maps were prepared. The first series presents heat maps that capture the dental hygienist rate (i.e., number of dental hygienists per 100,000 population) for all 520 FSAs included in the analysis. The second series presents standard heat maps that capture the dental hygienist rate for the 50 FSA-2s. Given the geographic size of Ontario (roughly equivalent to the combined size of Texas and New Mexico or the combined size of France and Spain) and the varying geographic size of the FSAs/FSA-2s, the heat maps tend to over-emphasize the geographically large FSAs/FSA-2s (typically rural and remote regions) and de-emphasize the geographically small FSAs/FSA-2s (typically urban areas). Therefore, each series presents an initial map of Ontario and then provides five additional heat maps that zoom into each of the five postal districts in the province (i.e., K, L, M, N, P) (see **Additional file 8**). As noted earlier, FSAs were originally defined to support postal sorting and delivery and not geospatial analysis, thus heat maps display some non-contiguous areas, particularly for the FSA-2 level maps, that can appear as small isolated ‘islands’ on the maps.

Visual analysis of the standard heat maps at the FSA level provides a fairly consistent picture of the variation in the dental hygienist rate (**Additional file 8: Heat Maps 2-7**). There is considerable variation across the province with pockets of high dental hygienist rates mostly in urban areas and the lowest dental hygienist rates across rural and remote areas of the province. Visual analysis of the standard heat maps at the FSA-2 level reveals distinct and larger geographic areas with the number of boundary regions reduced from 520 to 50 (**Additional file 8: Heat Maps 8-13**). This leads to less overall variation (reflected in the tighter scale) and helps to highlight the large rural/remote areas of low dental hygienist rates and the pockets of high dental hygienist rate FSA-2s. In particular, what is distinct from the FSA heat maps are large visible FSA-2s in four of the five postal districts (K, L, N, P). As noted earlier in the report, Canada Post designates rural/remote areas by a zero for the second character of the FSA, thus ‘K0’, ‘L0’, ‘N0’ and ‘P0’ occupy the majority of the visual space on the FSA-2 heat maps as they are capturing the rural/remote FSAs collectively. The M district, representing metropolitan Toronto, does not have an ‘M0’ FSA-2 in our dataset, therefore represents an exception.

Two interactive heat maps (at FSA and FSA-2 levels) were also generated to supplement the standard heat maps. The interactive heat maps are based on the same data but allow the viewer to zoom in/out and select and identify any FSA or FSA-2 region on either map to reveal its Canada Post location descriptor (for FSAs only) and specific dental hygienist rate.

## 4. Discussion

This project examined two linked datasets to assess the distribution of the dental hygienist workforce in Ontario. Both the Statistics Canada census dataset and the CDHO dental hygienist registrant dataset were of good quality and the data linkage was robust, with minimal data lost. Ultimately, 520 of a possible 532 FSAs and 13,843 of a possible 13,856 eligible dental hygienists in Ontario were examined.

Analyses of the two datasets were conducted both independently and as a linked dataset. Assessments were conducted at three levels of analysis, including for 520 FSAs, 50 FSA-2s and five FSA-1s. Twelve standard heat maps (six at the FSA level and six at the FSA-2 level) and two interactive (HTML) heat maps (one at the FSA level and one at the FSA-2 level) were produced.

The initial analyses revealed considerable and consistent variation among FSAs in terms of both the population count and the number of dental hygienists. The FSAs were not designed to have equal populations and this level of variation (from a low population of 5 to a high population of 115,850) provided the key base from which to analyze the distribution of dental hygienists in Ontario (which ranged from a low of zero to a high of 137). If there was perfect correlation between population and dental hygienist counts, one would expect the FSA data to show alignment between the two variables, irrespective of differing population counts. However, comparison of the distribution of the two variables by FSA suggests they were not particularly well-aligned. Furthermore, our analysis of the linked dataset, which focused on the dental hygienist rate (i.e., the number of dental hygienists per 100,000 population) indicated persistent variation, with FSA population count not a good predictor of the number of dental hygienists.

Overall, the dental hygienist rate across all 520 FSAs ranged from **zero to 20,000** dental hygienists per 100,000 population (or **zero to 739** if five extreme outlier FSAs are removed). Even when the dental hygienist rate was analyzed at increasing levels of aggregation, variable results were observed, albeit with an expected narrowing of the range:

- The dental hygienist rate ranged from **50 to 146** per 100,000 population for 50 FSA-2s
- The dental hygienist rate ranged from **91 to 101** per 100,000 population for 5 FSA-1s
- The dental hygienist rate was **97** per 100,000 population for the entire province of Ontario (13,843 dental hygienists for 14,223,936 people).

Based on these results, including considerable observed variation and an overall provincial dental hygienist rate of 97 dental hygienists per 100,000 population, it is not clear what an appropriate target rate should be. The calculated rate is somewhat higher than the 2021 rate published by the Canadian Institute for Health Information as part of its health workforce analysis (https://www.cihi.ca/sites/default/files/document/health-workforce-canada-2017-2021-overview-data-tables-en.xlsx). In that analysis, Ontario’s dental hygienist rate of 92 was the highest of any province or territory in Canada, which ranged from 52 to 92 dental hygienists per 100,000 population. However, based on the analysis presented here, 289 FSAs have a dental hygienist rate below 97, and 228 FSAs have a dental hygienist rate above 97.

Rather than determining a target provincial dental hygienist rate, it may be more valuable to understand the causes of the variability in the rate. Through these analyses, several FSAs were identified that could be targeted for more in-depth study. For example, 27 FSAs (K1X, K2R, K4C, K6K, K6T, K8B, K8R, L0H, L0N, L1Y, L3L, N1C, N1P, N3E, N3P, N3V, N4V, N4Z, N7W, N7X, P0G, P0Y, P1C, P3G, P3L, P7J, P7L) had no dental hygienists in the CDHO registrant database, while their respective populations ranged from 96 to 8,542 persons. Also, only five of the 520 FSAs (K1A, K1P, L5T, L5S, L4V) had dental hygienist rates per 100,000 population above 750, thus representing outliers that should be investigated further. There were also a few FSAs in urban areas that had low dental hygienist rates (e.g., L5E, L8L, N6L) or FSAs in rural areas with high dental hygienist rates (e.g.,

N4N) that conflicted with the broader pattern of results and thus represent targets for further investigation. It is also unclear to what extent the variation in the distribution of dental hygienists relates to the distribution of dentists in Ontario; however, it is important to note that, since 2007, regulatory changes have allowed patients direct access to dental hygienists without requiring a referral from a dentist. As a result, it is plausible that some regions may have differing rates of dentists and dental hygienists, particularly in rural and remote areas, where dental hygienists specializing in mobile services may provide greater accessibility compared to dentists.

### 4.1 Implications

Overall, the findings highlight stark disparities in the geographic distribution of dental hygienists across Ontario, underscoring the need for targeted policy and regulatory interventions. Based on these results, the following actions to address workforce imbalances are recommended:

- **Financial Incentives:** Health system stakeholders could offer financial incentives, such as rural practice grants, loan forgiveness programs, or reduced registration fees, to encourage dental hygienists to practice in underserved areas.
- **Policy Interventions:** Collaborative efforts between provincial policy-makers and local governments could integrate dental hygiene services into broader public health initiatives. For instance, mobile dental clinics and school-based oral health programs could be expanded to improve access in rural and remote communities.
- **Education and Training:** Incorporating rural placements and community-based training into dental hygiene education programs could expose students to the unique needs of underserved areas and foster interest in practicing in these communities.
- **Monitoring and Evaluation:** Regular geospatial analyses, like the one conducted in this study, could be institutionalized to monitor workforce trends, evaluate the impact of policy interventions, and identify emerging gaps in care.

### 4.2 Limitations and Considerations

As with any analysis, it is important to consider methodological issues that may have impacted the results and the interpretation of the findings. Four methodological considerations are highlighted, including (1) data quality and robustness of data linkage, (2) implications and value of multiple levels of analysis, (3) temporal issues, and (4) potential misalignment of oral health care seeking behaviours and census location data.

First, it should be noted that the analyses are dependent on the quality and accuracy of the postal code data contained in the two datasets. The Statistics Canada 2021 census dataset is a standardized and highly reliable data source, and the CDHO registrant dataset was found to be of reasonably high quality, with minimal concerns noted regarding the various fields included. The CDHO dataset contained only a few records with incomplete postal code information or non-Ontario postal codes. Where possible, included address information was used to identify complete postal codes and any records with non-Ontario postal codes were excluded. Thus, the geo-coding methods were valid and effective. Most date fields were also appropriately captured in the CDHO registrant dataset, however, a small handful of the dental hygienists in the database with an active membership status were older than would be expected (e.g., over 70 years). Given the small numbers, they were not excluded from the analysis.

Second, it is important to consider the implications of using three levels of analysis based on the FSA. From the outset, the main intention was to conduct the geospatial analysis at the FSA level. However, with the geographic coordinates in the boundary file assigned to the FSA, it was possible to aggregate the FSA data to a two-character FSA (FSA-2) level or a one-character FSA (FSA-1) level while maintaining the necessary geographic information. This allowed examination of the available data by 520 FSAs, 50 FSA-2s and 5 FSA-1s.

The 520 FSAs provide a large number of geographic units from which to conduct a basic assessment of the distribution of the dental hygienist workforce, but it does not necessarily represent logical geographic units for CDHO or its stakeholders. In contrast, the FSA-1 level of analysis provides very high-level insights across the five postal districts in Ontario (K, L, M, N, P), that are based on logical but also extremely large geographic areas. The FSA-2 level of analysis was initially predicted to offer a reasonable middle ground, yielding a reasonable number of geographic units (50) to conduct comparative analyses. However, the FSA-2s are impacted by a prevalence of non-contiguous geographic areas, which makes interpreting findings challenging. For example, a dental hygienist rate for an FSA-2 might be applicable to five or more non-contiguous areas, some not even remotely close to one another.

Third, it is important to note a few temporal considerations. The CDHO dataset was the most current of the two datasets used, with data drawn from the CDHO registrant database in May 2023. The census dataset was the most recent available but nevertheless was captured in 2021. Thus, there is a temporal discrepancy between the two datasets. An alternative approach would have been to acquire an older version of the CDHO data set (e.g., circa 2021), however, that would not take advantage of the most up-to-date registrant data. The 2021 census dataset is the most recent, comprehensive, and reliable population dataset available and while it is not perfectly aligned temporally with the CDHO dataset, it provides a reasonable proxy for the population distribution in 2023. It is acknowledged that cross-sectional data only allow assessment of the distribution of the dental hygienist workforce at a specific point in time and do not allow assessment of the changes in this workforce distribution over time. To address these temporal issues, repeating this type of geospatial analysis at regular intervals is recommended to be able to track changes in the distribution of the dental hygienist workforce longitudinally. It may also make sense to consider aligning future geospatial analyses with upcoming releases of new census data to achieve a closer temporal match.

Fourth, given the datasets used, there is a potential misalignment between individual oral health care seeking behaviours and census location data. The dental hygienist postal code information is linked to the dental hygienist’s primary work location, while the census data is based on an individual’s home address. Therefore, this does not adequately account for situations where a patient may be more likely to seek dental hygienist services closer to their location of work than their home. Given the upheaval in work/home balances due to the COVID-19 pandemic, the resultant increases in remote working, and the increasing availability of mobile dental hygienist services, it is likely that the trend is moving towards people seeking oral health services closer to their home than workplace locations. Nevertheless, it is clear that we are still in the midst of a period of substantive change in this regard, therefore, this limitation is not something that can be easily solved, even if postal code data on both work and home locations were publicly available. However, this geospatial analysis provides a reasonable assessment of the distribution of the dental hygienist workforce relative to the prevailing population.

## 5. Conclusions

The application of geographic information and spatial analysis methods in this study has yielded comprehensive insights into the distribution of the dental hygienist workforce in Ontario across multiple levels of analysis. These findings reveal significant geographic disparities in workforce distribution, with implications for planning and policy. To address these challenges, actionable strategies are proposed, including financial incentives for rural practice, integration of dental hygiene into public health frameworks, and community-based training programs. These recommendations, alongside continued geospatial monitoring, can guide efforts to improve the alignment of workforce distribution with population needs and ensure access to oral health care across Ontario.

Furthermore, the rigorous methodological approach and thorough evaluation of potential limitations offer valuable guidance for researchers conducting similar analyses for other jurisdictions. These findings are important for informing discussions, regulatory actions, and policy decisions among key stakeholders, not only within Ontario but also in other jurisdictions facing similar workforce distribution challenges. By providing a detailed framework and insights that extend beyond Ontario, this study contributes to the global understanding of dental hygienist workforce dynamics and supports the development of informed policies on a broader scale.

## Data Availability

All data generated or analyzed during this study are included in this published article (and its Additional files 1-8).

## List of Abbreviations

CDHO: College of Dental Hygienists of Ontario
CIHI: Canadian Institute of Health Information
COVID-19: Coronavirus Disease 2019
FSA: Forward Sortation Area
GIS: Geographic Information Systems
HTML: HyperText Markup Language

## Declarations

### Clinical trial number

Not applicable.

### Ethics approval and consent to participate

Not Applicable. The University of Toronto Research Ethics Board exempts research from human ethics review under certain situations, including when the research “…relies exclusively on information that is either of the following: (a) Publicly available through a mechanism set out by legislation or regulation and that is protected by law (e.g. Statistics Canada files), (b) In the public domain and the individuals to whom the information refers have no reasonable expectation of privacy (non-intrusive, does not involve direct interaction between the researcher and individuals through the Internet)” (https://research.utoronto.ca/ethics-human-research/activities-exempt-human-ethics-review). As our geospatial analysis relies on publicly available Statistics Canada census data and CDHO registrant data, this study did not require research ethics board approval.

### Consent for publication

Not applicable.

### Authors’ contributions

Conceptualization: MJD, GP; analysis/methodology design: MJD; registrant data collection: EB, KS, GP; population data collection: MJD; data analysis: MJD; preparation of initial draft of manuscript: MJD; reviewing/revising manuscript: MJD, EB, KS, GP. All authors read and approved the submitted version of the manuscript.

## Acknowledgements

The authors thank the College of Dental Hygienists of Ontario for providing access to an anonymized registrant dataset to facilitate the geospatial analysis.

## Authors’ information

^1^Institute of Health Policy, Management and Evaluation, Dalla Lana School of Public Health, University of Toronto, Toronto, Ontario, Canada: MJD

^2^Accessing Centre for Expertise, Toronto, Ontario, Canada: MJD

^3^College of Dental Hygienists of Ontario, Toronto, Ontario, Canada: EB, KS, GP

