## Additional Files for "Comparison of the Distribution of the Dental Hygienist Workforce and Population in Ontario: A Geospatial Analysis"

### MedRxiv Preprint

### Title Page

[1]Institute of Health Policy, Management and Evaluation
Dalla Lana School of Public Health
University of Toronto
155 College Street, Suite 425
Toronto, ON M5T 3M6

[2]Accessing Centre for Expertise
155 College Street, Suite 425
Toronto, ON M5T 3M6

[3]College of Dental Hygienists of Ontario
175 Bloor Street East
North Tower, Suite 601
Toronto, ON M4W 3R8

*Corresponding author

**Keywords**
Dental hygienist; workforce; supply; demand; scoping review

#### Additional File 1: List of Ontario Forward Sortation Areas (FSAs) and Canada Post Location Descriptors[27]

| FSA | Canada Post Location Descriptors | FSA | Canada Post Location Descriptors | FSA | Canada Post Location Descriptors | FSA | Canada Post Location Descriptors |
|---|---|---|---|---|---|---|---|
| K0A | OTTAWA ON DCF | K8B | PEMBROKE ON LCD MAIN | L3X | NEWMARKET ON LCD 1 | L7T | BURLINGTON ON LCD 1 |
| K0B | OTT EXT-HAWKESBURY ON DCF | K8H | PETAWAWA ON LCD MAIN | L3Y | NEWMARKET ON LCD 1 | L8B | WATERDOWN ON STN WATERDOWN |
| K0C | OTT EXT-CORNWALL ON DCF | K8N | BELLEVILLE ON STN MAIN | L3Z | BRADFORD ON LCD MAIN | L8E | STONEY CREEK ON STN LCD 3 |
| K0E | OTT EXT-BROCKVILLE ON DCF | K8P | BELLEVILLE ON STN MAIN | L4A | STOUFFVILLE ON LCD MAIN | L8G | STONEY CREEK ON STN LCD 3 |
| K0G | OTT EXT-SMITHS FALLS ON DCF | K8R | BELLEVILLE ON STN MAIN | L4B | RICHMOND HILL ON LCD 20 | L8H | STONEY CREEK ON STN LCD 3 |
| K0H | TOR EXT-KINGSTON ON DCF | K8V | TRENTON ON LCD MAIN | L4C | RICHMOND HILL ON STN A | L8J | HAMILTON ON STN LCD 5 |
| K0J | OTT EXT-PEMBROKE ON DCF | K9A | COBOURG ON LCD MAIN | L4E | RICHMOND HILL ON LCD 14 | L8K | STONEY CREEK ON STN LCD 3 |
| K0K | TOR EXT-BELLEVILLE ON DCF | K9H | PETERBOROUGH ON STN DELIVERY | L4G | AURORA ON LCD MAIN | L8L | HAMILTON ON STN LCD 1 |
| K0L | TOR EXT-PETERBOROUGH ON DCF | K9J | PETERBOROUGH ON STN DELIVERY | L4H | WOODBRIDGE ON STN MAIN | L8M | HAMILTON ON STN LCD 1 |
| K0M | TOR EXT-PETERBOROUGH ON DCF | K9K | PETERBOROUGH ON STN DELIVERY | L4J | THORNHILL ON LCD MAIN | L8N | HAMILTON ON STN LCD 1 |
| K1A | OTTAWA ON STN FED-GOVT/GOUV | K9L | PETERBOROUGH ON STN DELIVERY | L4K | CONCORD ON STN MAIN | L8P | HAMILTON ON STN LCD 1 |
| K1B | OTTAWA ON LCD LANCASTER | K9V | LINDSAY ON LCD MAIN | L4L | WOODBRIDGE ON STN MAIN | L8R | HAMILTON ON STN LCD 1 |
| K1C | ORLEANS ON STN ORLEANS | | | L4M | BARRIE ON LCD DISTRIBUTION | L8S | HAMILTON ON STN LCD 1 |
| K1E | ORLEANS ON STN ORLEANS | L0A | TORONTO WEST # 2 ON DCF | L4N | BARRIE ON LCD DISTRIBUTION | L8T | HAMILTON ON STN LCD 5 |
| K1G | OTTAWA ON LCD LANCASTER | L0B | TORONTO WEST # 2 ON DCF | L4P | KESWICK ON LCD MAIN | L8V | HAMILTON ON STN LCD 5 |
| K1H | OTTAWA ON LCD LANCASTER | L0C | TOR EXT-NEWMARKET ON DCF | L4R | MIDLAND ON LCD MAIN | L8W | HAMILTON ON STN LCD 5 |
| K1J | OTTAWA ON STN TERMINAL 2 | L0E | TOR EXT-NEWMARKET ON DCF | L4S | RICHMOND HILL ON LCD 14 | L9A | HAMILTON ON STN LCD 5 |
| K1K | OTTAWA ON STN TERMINAL 2 | L0G | TOR EXT-NEWMARKET ON DCF | L4T | MISSISSAUGA ON LCD MALTON | L9B | HAMILTON ON LCD WEST |
| K1L | OTTAWA ON STN VANIER | L0H | TORONTO WEST # 2 ON DCF | L4V | MISSISSAUGA ON LCD MALTON | L9C | HAMILTON ON STN LCD 5 |
| K1M | OTTAWA ON STN VANIER | L0J | TORONTO WEST # 2 ON DCF | L4W | MISSISSAUGA ON LCD 2 | L9E | MILTON ON LCD MAIN |
| K1N | OTTAWA ON STN VANIER | L0K | BARRIE ON DCF | L4X | MISSISSAUGA ON LCD 2 | L9G | HAMILTON ON LCD WEST |
| K1P | OTTAWA ON LCD S. FLEMING | L0L | BARRIE ON DCF | L4Y | MISSISSAUGA ON LCD 2 | L9H | HAMILTON ON LCD WEST |
| K1R | OTTAWA ON LCD S. FLEMING | L0M | BARRIE ON DCF | L4Z | MISSISSAUGA ON LCD 5 | L9J | BARRIE ON LCD DISTRIBUTION |
| K1S | OTTAWA ON LCD S. FLEMING | L0N | TORONTO WEST # 2 ON DCF | L5A | MISSISSAUGA ON LCD 5 | L9K | HAMILTON ON LCD WEST |
| K1T | OTTAWA ON LCD S | L0P | TORONTO WEST # 2 ON DCF | L5B | MISSISSAUGA ON LCD 5 | L9L | PORT PERRY ON LCD MAIN |
| K1V | OTTAWA ON LCD S | L0R | HAMILTON ON DCF | L5C | MISSISSAUGA ON STN LCD 3 | L9M | PENETANGUISHENE ON LCD MAIN |
| K1W | ORLEANS ON STN ORLEANS | L0S | HAMILTON ON DCF | L5E | MISSISSAUGA ON STN PORT CREDIT | L9N | EAST GWILLIMBURY ON LCD MAIN |
| K1X | OTTAWA ON LCD LANCASTER | L0V†‡ | OTTAWA ON DCF | L5G | MISSISSAUGA ON STN PORT CREDIT | L9P | UXBRIDGE ON STN MAIN |
| K1Y | OTTAWA ON STN C | L1A | PORT HOPE ON LCD MAIN | L5H | MISSISSAUGA ON STN PORT CREDIT | L9R | ALLISTON ON LCD MAIN |
| K1Z | OTTAWA ON STN C | L1B | BOWMANVILLE ON LCD MAIN | L5J | MISSISSAUGA ON STN PORT CREDIT | L9S | INNISFIL ON LCD MAIN |
| K2A | OTTAWA ON STN J | L1C | BOWMANVILLE ON LCD MAIN | L5K | MISSISSAUGA ON STN LCD 3 | L9T | MILTON ON LCD MAIN |
| K2B | OTTAWA ON STN J | L1E | BOWMANVILLE ON LCD MAIN | L5L | MISSISSAUGA ON STN LCD 3 | L9V | SHELBURNE ON LCD MAIN |
| K2C | NEPEAN ON LCD MERIVALE | L1G | OSHAWA ON STN A | L5M | MISSISSAUGA ON LCD 4 | L9W | ORANGEVILLE ON LCD MAIN |
| K2E | NEPEAN ON LCD MERIVALE | L1H | OSHAWA ON STN A | L5N | MISSISSAUGA ON LCD 4 | L9X | BARRIE ON LCD DISTRIBUTION |
| K2G | NEPEAN ON LCD MERIVALE | L1J | OSHAWA ON STN A | L5P†‡ | MISSISSAUGA ON STN TORONTO AMF | L9Y | COLLINGWOOD ON LCD |
| K2H | NEPEAN ON STN H | L1K | OSHAWA ON STN A | L5R | MISSISSAUGA ON LCD 5 | L9Z | WASAGA BEACH ON LCD MAIN |
| K2J | NEPEAN ON LCD N | L1L | OSHAWA ON STN A | L5S | MISSISSAUGA ON LCD MALTON | | |
| K2K | KANATA ON STN KANATA | L1M | WHITBY ON LCD BROOKLIN | L5T | MISSISSAUGA ON LCD MALTON | M0B†‡ | TORONTO WEST # 2 ON DCF |
| K2L | KANATA ON STN KANATA | L1N | WHITBY ON LCD WHITBY | L5V | MISSISSAUGA ON LCD 4 | M1B | TORONTO ON LCD MALVERN |
| K2M | KANATA ON STN KANATA | L1P | WHITBY ON LCD WHITBY | L5W | MISSISSAUGA ON LCD 4 | M1C | TORONTO ON LCD WEST HILL |
| K2P | OTTAWA ON LCD S. FLEMING | L1R | WHITBY ON LCD WHITBY | L6A | MAPLE ON STN DELIVERY CENTRE | M1E | TORONTO ON LCD WEST HILL |
| K2R | NEPEAN ON LCD N | L1S | AJAX ON STN DELIVERY CENTRE | L6B | UNIONVILLE ON LCD 2 | M1G | SCARBOROUGH ON LCD C |
| K2S | STITTSVILLE ON STN B | L1T | AJAX ON STN DELIVERY CENTRE | L6C | UNIONVILLE ON LCD 2 | M1H | SCARBOROUGH ON LCD C |
| K2T | KANATA ON STN KANATA | L1V | PICKERING ON LCD MAIN | L6E | UNIONVILLE ON LCD 2 | M1J | SCARBOROUGH ON LCD 11 |
| K2V | KANATA ON STN KANATA | L1W | PICKERING ON LCD MAIN | L6G | UNIONVILLE ON LCD 1 | M1K | SCARBOROUGH ON LCD 11 |
| K2W | KANATA ON STN KANATA | L1X | PICKERING ON LCD MAIN | L6H | OAKVILLE ON STN MAIN | M1L | SCARBOROUGH ON LCD 11 |
| K4A | ORLEANS ON STN ORLEANS | L1Y | PICKERING ON LCD MAIN | L6J | OAKVILLE ON STN MAIN | M1M | SCARBOROUGH ON LCD 11 |
| K4B | NAVAN ON STN MAIN | L1Z | AJAX ON STN DELIVERY CENTRE | L6K | OAKVILLE ON LCD 1 | M1N | SCARBOROUGH ON LCD 11 |
| K4C | CUMBERLAND ON STN MAIN | L2A | FORT ERIE ON LCD MAIN | L6L | OAKVILLE ON LCD 1 | M1P | SCARBOROUGH ON LCD D |
| K4K | ROCKLAND ON STN MAIN | L2E | NIAGARA FALLS ON LCD MAIN | L6M | OAKVILLE ON LCD 1 | M1R | SCARBOROUGH ON LCD D |
| K4M | MANOTICK ON STN MAIN | L2G | NIAGARA FALLS ON LCD MAIN | L6P | BRAMPTON ON LCD 4 | M1S | TORONTO ON LCD AGINCOURT |
| K4P | MANOTICK ON STN MAIN | L2H | NIAGARA FALLS ON LCD MAIN | L6R | BRAMPTON ON LCD 4 | M1T | TORONTO ON LCD AGINCOURT |
| K4R | RUSSELL ON STN MAIN | L2J | NIAGARA FALLS ON LCD MAIN | L6S | BRAMPTON ON LCD B | M1V | TORONTO ON LCD AGINCOURT 4 |
| K6A | HAWKESBURY ON LCD MAIN | L2M | ST CATHARINES ON STN LCD 1 | L6T | BRAMPTON ON LCD B | M1W | SCARBOROUGH ON STN F |
| K6H | CORNWALL ON LCD 1 | L2N | ST CATHARINES ON STN LCD 1 | L6V | BRAMPTON ON LCD 1 | M1X | TORONTO ON LCD AGINCOURT 4 |
| K6J | CORNWALL ON LCD 1 | L2P | ST CATHARINES ON LCD 2 | L6W | BRAMPTON ON LCD B | M2H | WILLOWDALE ON LCD 2 |
| K6K | CORNWALL ON LCD 1 | L2R | ST CATHARINES ON LCD 2 | L6X | BRAMPTON ON LCD 1 | M2J | SCARBOROUGH ON STN F |
| K6T | BROCKVILLE ON LCD MAIN | L2S | ST CATHARINES ON LCD 2 | L6Y | BRAMPTON ON LCD B | M2K | WILLOWDALE ON LCD B |
| K6V | BROCKVILLE ON LCD MAIN | L2T | ST CATHARINES ON LCD 2 | L6Z | BRAMPTON ON LCD 1 | M2L | WILLOWDALE ON LCD L |
| K7A | SMITHS FALLS ON LCD MAIN | L2V | ST CATHARINES ON LCD 2 | L7A | BRAMPTON ON LCD 1 | M2M | WILLOWDALE ON LCD 2 |
| K7C | CARLETON PLACE ON LCD MAIN | L2W | ST CATHARINES ON STN LCD 1 | L7B | KING CITY ON STN MAIN | M2N | WILLOWDALE ON LCD N |
| K7G | GANANOQUE ON LCD MAIN | L3B | WELLAND ON LCD MAIN | L7C | CALEDON ON STN MAIN | M2P | WILLOWDALE ON LCD L |
| K7H | PERTH ON STN MAIN | L3C | WELLAND ON LCD MAIN | L7E | CALEDON ON STN MAIN | M2R | WILLOWDALE ON STN D |
| K7K | KINGSTON ON LCD MAIN | L3J§ | BEAMSVILLE ON LCD MAIN | L7G | GEORGETOWN ON LCD MAIN | M3A | TORONTO ON LCD DON MILLS |
| K7L | KINGSTON ON LCD MAIN | L3K | PORT COLBORNE ON LCD MAIN | L7J | ACTON ON LCD MAIN | M3B | TORONTO ON LCD DON MILLS |
| K7M | KINGSTON ON STN A | L3L | WOODBRIDGE ON STN MAIN | L7K | CALEDON ON STN C | M3C | TORONTO ON LCD DON MILLS |
| K7N | KINGSTON ON STN A | L3M | GRIMSBY ON LCD MAIN | L7L | BURLINGTON ON LCD 1 | M3H | TORONTO ON LCD DOWNSVIEW A |
| K7P | KINGSTON ON STN A | L3P | UNIONVILLE ON LCD 1 | L7M | BURLINGTON ON LCD 1 | M3J | TORONTO ON LCD DOWNSVIEW C |

| FSA | Canada Post Location Descriptors | FSA | Canada Post Location Descriptors | FSA | Canada Post Location Descriptors | FSA | Canada Post Location Descriptors |
|---|---|---|---|---|---|---|---|
| K7R | NAPANEE ON LCD MAIN | L3R | UNIONVILLE ON LCD 1 | L7N | BURLINGTON ON LCD 1 | M3K | TORONTO ON LCD DOWNSVIEW A |
| K7S | ARNPRIOR ON LCD MAIN | L3S | UNIONVILLE ON LCD 2 | L7P | BURLINGTON ON LCD 1 | M3L | TORONTO ON LCD DOWNSVIEW A |
| K7V | RENFREW ON LCD MAIN | L3T | THORNHILL ON LCD 20 | L7R | BURLINGTON ON LCD 1 | M3M | TORONTO ON LCD DOWNSVIEW A |
| K8A | PEMBROKE ON LCD MAIN | L3V | ORILLIA ON LCD MAIN | L7S | BURLINGTON ON LCD 1 | M3N | TORONTO ON LCD DOWNSVIEW C |
| M4A | NORTH YORK ON STN O | M9L | TORONTO ON LCD WESTON B | N4B | DELHI ON LCD MAIN | N9K | WINDSOR ON LCD TECUMSEH |
| M4B | NORTH YORK ON STN O | M9M | TORONTO ON LCD WESTON B | N4G | TILLSONBURG ON LCD MAIN | N9V | AMHERSTBURG ON LCD MAIN |
| M4C | TORONTO ON STN H | M9N | YORK ON STN WESTON A | N4K | OWEN SOUND ON LCD MAIN | N9Y | KINGSVILLE ON LCD MAIN |
| M4E | TORONTO ON STN H | M9P | YORK ON STN WESTON A | N4L | MEAFORD ON STN MAIN | | |
| M4G | TORONTO ON STN R | M9R | YORK ON STN WESTON A | N4N | HANOVER ON LCD MAIN | P0A | BARRIE ON DCF |
| M4H | TORONTO ON STN R | M9V | ETOBICOKE ON STN C | N4S | WOODSTOCK ON LCD MAIN | P0B | BARRIE ON DCF |
| M4J | TORONTO ON STN J | M9W | ETOBICOKE ON LCD B | N4T | WOODSTOCK ON LCD MAIN | P0C | BARRIE ON DCF |
| M4K | TORONTO ON STN J | | | N4V | WOODSTOCK ON LCD MAIN | P0E | BARRIE ON DCF |
| M4L | TORONTO ON STN G | N0A | HAMILTON ON DCF | N4W | LISTOWEL ON LCD MAIN | P0G | BARRIE ON DCF |
| M4M | TORONTO ON STN G | N0B | KITCHENER ON DCF | N4X | ST MARYS ON STN MAIN | P0H | NORTH BAY ON DCF |
| M4N | TORONTO ON LCD K | N0C | OWEN SOUND ON DCF | N4Z | STRATFORD ON LCD MAIN | P0J | SUDBURY ON DCF |
| M4P | TORONTO ON LCD K | N0E | HAMILTON ON DCF | N5A | STRATFORD ON LCD MAIN | P0K | SUDBURY ON DCF |
| M4R | TORONTO ON LCD K | N0G | KITCHENER ON DCF | N5C | INGERSOLL ON LCD MAIN | P0L | TIMMINS ON DCF |
| M4S | TORONTO ON LCD Q | N0H | OWEN SOUND ON DCF | N5H | AYLMER ON LCD MAIN | P0M | SUDBURY ON DCF |
| M4T | TORONTO ON LCD Q | N0J | LONDON ON DCF | N5L | PORT STANLEY ON STN MAIN | P0N | TIMMINS ON DCF |
| M4V | TORONTO ON LCD Q | N0K | KITCHENER ON DCF | N5P | ST THOMAS ON LCD MAIN | P0P | SUDBURY ON DCF |
| M4W | TORONTO ON LCD F | N0L | LONDON ON DCF | N5R | ST THOMAS ON LCD MAIN | P0R | SUDBURY ON DCF |
| M4X | TORONTO ON LCD ADELAIDE | N0M | LONDON ON DCF | N5V | LONDON ON LCD 1 | P0S | SAULT STE. MARIE ON DCF |
| M4Y | TORONTO ON LCD CHARLES ST | N0N | LONDON ON DCF | N5W | LONDON ON LCD 1 | P0T | THUNDER BAY ON DCF |
| M5A | TORONTO ON LCD ADELAIDE | N0P | LONDON ON DCF | N5X | LONDON ON LCD 1 | P0V | DRYDEN ON DCF |
| M5B | TORONTO ON LCD 2 | N0R | WINDSOR ON DCF | N5Y | LONDON ON LCD 1 | P0W | DRYDEN ON DCF |
| M5C | TORONTO ON LCD 2 | N1A | DUNNVILLE ON LCD MAIN | N5Z | LONDON ON LCD 6 | P0X | DRYDEN ON DCF |
| M5E | TORONTO ON LCD 2 | N1C | GUELPH ON LCD ROYAL CITY MAIL | N6A | LONDON ON LCD 6 | P0Y | WINNIPEG # 2 MB DCF |
| M5G | TORONTO ON LCD 2 | N1E | GUELPH ON LCD ROYAL CITY MAIL | N6B | LONDON ON LCD 6 | P1A | NORTH BAY ON LCD MAIN |
| M5H | TORONTO ON LCD 1 | N1G | GUELPH ON LCD ROYAL CITY MAIL | N6C | LONDON ON LCD 5 | P1B | NORTH BAY ON LCD MAIN |
| M5J | TORONTO ON LCD 1 | N1H | GUELPH ON LCD ROYAL CITY MAIL | N6E | LONDON ON LCD 5 | P1C | NORTH BAY ON LCD MAIN |
| M5K†‡ | TORONTO ON STN TORONTO DOM | N1K | GUELPH ON LCD ROYAL CITY MAIL | N6G | LONDON ON LCD 4 | P1H | HUNTSVILLE ON STN MAIN |
| M5L†‡ | TORONTO ON STN COMMERCE COURT | N1L | GUELPH ON LCD ROYAL CITY MAIL | N6H | LONDON ON LCD 4 | P1L | BRACEBRIDGE ON LCD MAIN |
| M5M | TORONTO ON STN S | N1M | FERGUS ON LCD MAIN | N6J | LONDON ON LCD 3 | P1P | GRAVENHURST ON STN MAIN |
| M5N | TORONTO ON STN S | N1P | KITCHENER ON LCD GALT | N6K | LONDON ON LCD 3 | P2A | PARRY SOUND ON LCD MAIN |
| M5P | TORONTO ON STN S | N1R | KITCHENER ON LCD GALT | N6L | LONDON ON LCD 5 | P2B | STURGEON FALLS ON STN MAIN |
| M5R | TORONTO ON LCD P | N1S | KITCHENER ON LCD GALT | N6M | LONDON ON LCD 1 | P2N | KIRKLAND LAKE ON LCD MAIN |
| M5S | TORONTO ON LCD P | N1T | KITCHENER ON LCD GALT | N6N | LONDON ON LCD 5 | P3A | SUDBURY ON LCD MAIN |
| M5T | TORONTO ON LCD B | N2A | KITCHENER ON LCD 1 | N6P | LONDON ON STN LAMBETH | P3B | SUDBURY ON LCD MAIN |
| M5V | TORONTO ON LCD B | N2B | KITCHENER ON LCD 2 | N7A | GODERICH ON LCD MAIN | P3C | SUDBURY ON LCD MAIN |
| M5W†‡ | TORONTO ON STN A | N2C | KITCHENER ON LCD 1 | N7G | STRATHROY ON LCD MAIN | P3E | SUDBURY ON LCD MAIN |
| M5X†‡ | TORONTO ON STN 1ST CAN PLACE | N2E | KITCHENER ON LCD 3 | N7L | CHATHAM ON LCD MAIN | P3G | SUDBURY ON LCD MAIN |
| M6A | TORONTO ON STN T | N2G | KITCHENER ON LCD 2 | N7M | CHATHAM ON LCD MAIN | P3L | GARSON ON STN MAIN |
| M6B | TORONTO ON STN T | N2H | KITCHENER ON LCD 2 | N7S | SARNIA ON LCD MAIN | P3N | VAL CARON ON STN MAIN |
| M6C | TORONTO ON STN L | N2J | KITCHENER ON LCD WATERLOO | N7T | SARNIA ON LCD MAIN | P3P | HANMER ON STN MAIN |
| M6E | TORONTO ON STN L | N2K | KITCHENER ON LCD WATERLOO | N7V | SARNIA ON LCD MAIN | P3Y | LIVELY ON STN MAIN |
| M6G | TORONTO ON LCD E | N2L | KITCHENER ON LCD WATERLOO | N7W | SARNIA ON LCD MAIN | P4N | TIMMINS ON LCD MAIN |
| M6H | TORONTO ON LCD E | N2M | KITCHENER ON LCD 3 | N7X | SARNIA ON LCD MAIN | P4P | TIMMINS ON LCD MAIN |
| M6J | TORONTO ON LCD C | N2N | KITCHENER ON LCD 3 | N8A | WALLACEBURG ON LCD MAIN | P4R | TIMMINS ON LCD MAIN |
| M6K | TORONTO ON LCD C | N2P | KITCHENER ON LCD 3 | N8H | LEAMINGTON ON LCD MAIN | P5A | ELLIOT LAKE ON LCD MAIN |
| M6L | NORTH YORK ON STN W | N2R | KITCHENER ON LCD 3 | N8M | ESSEX ON LCD MAIN | P5E | ESPANOLA ON STN MAIN |
| M6M | NORTH YORK ON STN W | N2T | KITCHENER ON LCD WATERLOO | N8N | WINDSOR ON LCD TECUMSEH | P5N | KAPUSKASING ON LCD MAIN |
| M6N | TORONTO ON STN L | N2V | KITCHENER ON LCD WATERLOO | N8P | WINDSOR ON LCD TECUMSEH | P6A | SAULT STE. MARIE ON LCD MAIN |
| M6P | TORONTO ON LCD 3 | N2Z | KINCARDINE ON STN MAIN | N8R | WINDSOR ON LCD 4 | P6B | SAULT STE. MARIE ON LCD MAIN |
| M6R | TORONTO ON LCD C | N3A | NEW HAMBURG ON STN MAIN | N8S | WINDSOR ON LCD 1 | P6C | SAULT STE. MARIE ON LCD MAIN |
| M6S | TORONTO ON LCD 3 | N3B | ELMIRA ON LCD MAIN | N8T | WINDSOR ON LCD 1 | P7A | THUNDER BAY ON STN P |
| M7A‡ | TORONTO ON STN PROVINCIAL GOVT | N3C | KITCHENER ON PRESTON | N8V†‡ | WINDSOR ON LCD 4 | P7B | THUNDER BAY ON STN P |
| M7R†‡ | MISSISSAUGA ON STN GATEWAY CR | N3E | KITCHENER ON PRESTON | N8W | WINDSOR ON LCD 4 | P7C | THUNDER BAY ON STN F |
| M7Y†‡ | TORONTO ON STN BRM B | N3H | KITCHENER ON PRESTON | N8X | WINDSOR ON LCD 3 | P7E | THUNDER BAY ON STN F |
| M8V | ETOBICOKE ON STN N | N3L | BRANTFORD ON LCD MAIN | N8Y | WINDSOR ON LCD 4 | P7G | THUNDER BAY ON STN P |
| M8W | ETOBICOKE ON STN N | N3P | BRANTFORD ON LCD MAIN | N9A | WINDSOR ON LCD 3 | P7J | THUNDER BAY ON STN F |
| M8X | ETOBICOKE ON LCD D | N3R | BRANTFORD ON LCD MAIN | N9B | WINDSOR ON LCD 3 | P7K | THUNDER BAY ON STN F |
| M8Y | ETOBICOKE ON LCD U | N3S | BRANTFORD ON LCD MAIN | N9C | WINDSOR ON LCD 3 | P7L | THUNDER BAY ON STN F |
| M8Z | ETOBICOKE ON LCD U | N3T | BRANTFORD ON LCD MAIN | N9E | WINDSOR ON LCD 2 | P8N | DRYDEN ON LCD MAIN |
| M9A | ETOBICOKE ON LCD D | N3V | BRANTFORD ON LCD MAIN | N9G | WINDSOR ON LCD 4 | P8T | SIOUX LOOKOUT ON STN MAIN |
| M9B | ETOBICOKE ON LCD D | N3W | CALEDONIA ON STN MAIN | N9H | WINDSOR ON LCD 4 | P9A | FORT FRANCES ON LCD MAIN |
| M9C | ETOBICOKE ON LCD A | N3Y | SIMCOE ON LCD MAIN | N9J | WINDSOR ON LCD 2 | P9N | KENORA ON LCD MAIN |

FSA – Forward Sortation Area
†FSA not included in analyses (zero population count in 2021 census dataset)
‡FSA not included in analyses (no geographic definition exists in Statistics Canada boundary shape file)
§FSA not included in analyses (Canada Post added new FSA in 2022 therefore not included in 2021 census dataset)

#### Additional File 2: FSAs Excluded from Analysis

| FSA | Dental Hygienists (CDHO dataset) | Population (census dataset) | Dental Hygienists per 100,000 Population Rate Calculable | Canada Post Location Descriptor (Additional Details) | Reason for Exclusion |
|---|---|---|---|---|---|
| L0V | 0 | 0 | No | OTTAWA ON DCF | Not included in census dataset (zero population) |
| L3J | 1 | 0 | No | BEAMSVILLE ON LCD MAIN | Not included in census dataset (new FSA - 2022) |
| L5P | 0 | 0 | No | MISSISSAUGA ON STN TORONTO AMF (YYZ-Pearson International Airport) | Not included in census dataset (zero population) |
| M0B | 0 | 0 | No | TORONTO WEST # 2 ON DCF | Not included in census dataset (zero population) |
| M5K | 3 | 0 | No | TORONTO ON STN TORONTO DOM (Toronto Dominion Building) | Not included in census dataset (zero population) |
| M5L | 0 | 0 | No | TORONTO ON STN COMMERCE COURT | Not included in census dataset (zero population) |
| M5W | 0 | 0 | No | TORONTO ON STN A | Not included in census dataset (zero population) |
| M5X | 8 | 0 | No | TORONTO ON STN 1ST CAN PLACE (Toronto First Canadian Place) | Not included in census dataset (zero population) |
| M7A | 1 | 5 | Yes | TORONTO ON STN PROVINCIAL GOVT (Queen’s Park) | No Statistics Canada boundary file available |
| M7R | 0 | 0 | No | MISSISSAUGA ON STN GATEWAY CR (Canada Post Gateway) | Not included in census dataset (zero population) |
| M7Y | 0 | 0 | No | TORONTO ON STN BRM B (Business Reply Mail Processing Centre) | Not included in census dataset (zero population) |
| N8V | 0 | 0 | No | WINDSOR ON LCD 4 (YQG – Windsor International Airport) | Not included in census dataset (zero population) |

FSA – Forward Sortation Area
CDHO – College of Dental Hygienists

#### Additional File 3: Population per Forward Sortation Area (FSA) in Ontario

##### Figure AF3a: Population per Forward Sortation Area (FSA) in Ontario

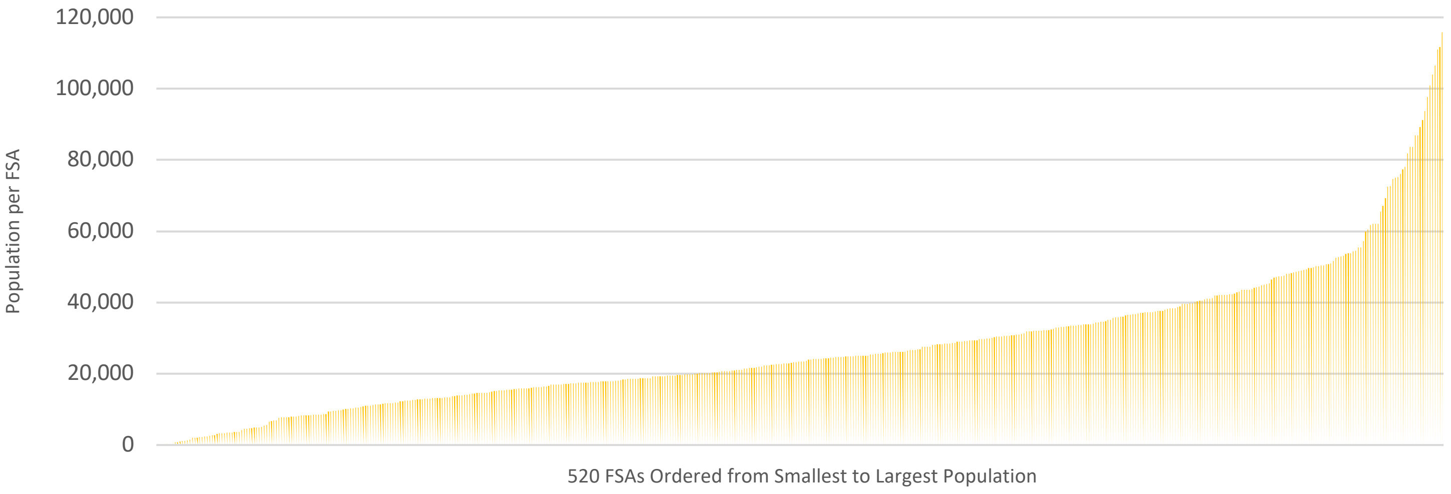


##### Figure AF3b: Population per Forward Sortation Area-2 (FSA-2) in Ontario

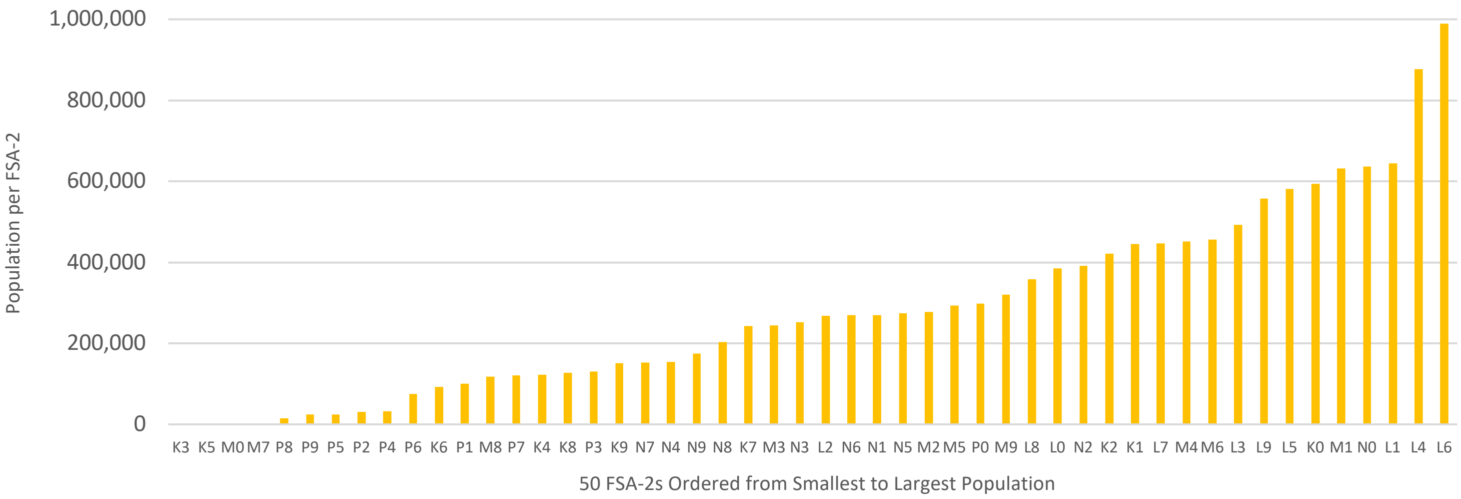


##### Figure AF3c: Population per Forward Sortation Area-1 (FSA-1) in Ontario

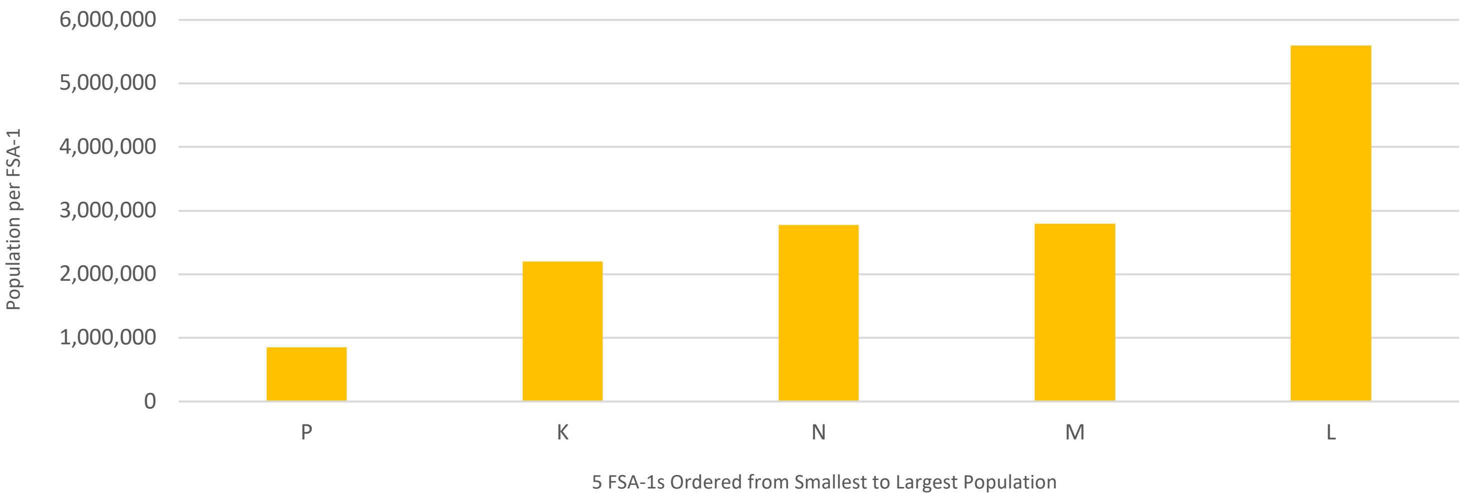

#### Additional File 4: Dental Hygienists, Population and Dental Hygienist Rate per 520 Forward Sortation Areas (FSAs)

| FSA | Dental Hygienists | Population | Dental Hygienists per 100,000 Population |
|---|---|---|---|
| K0A | 64 | 111,626 | 57 |
| K0B | 6 | 21,020 | 29 |
| K0C | 26 | 52,838 | 49 |
| K0E | 14 | 39,649 | 35 |
| K0G | 38 | 39,862 | 95 |
| K0H | 6 | 47,286 | 13 |
| K0J | 22 | 34,322 | 64 |
| K0K | 53 | 115,850 | 46 |
| K0L | 46 | 78,142 | 59 |
| K0M | 20 | 53,815 | 37 |
| K1A | 8 | 594 | 1,347 |
| K1B | 14 | 17,769 | 79 |
| K1C | 67 | 37,235 | 180 |
| K1E | 21 | 14,508 | 145 |
| K1G | 39 | 35,074 | 111 |
| K1H | 57 | 16,306 | 350 |
| K1J | 32 | 28,268 | 113 |
| K1K | 32 | 32,046 | 100 |
| K1L | 23 | 18,213 | 126 |
| K1M | 3 | 6,764 | 44 |
| K1N | 43 | 26,639 | 161 |
| K1P | 21 | 645 | 3,256 |
| K1R | 7 | 20,343 | 34 |
| K1S | 32 | 31,257 | 102 |
| K1T | 11 | 40,054 | 27 |
| K1V | 42 | 57,157 | 73 |
| K1W | 7 | 13,825 | 51 |
| K1X | 0 | 4,594 | - |
| K1Y | 42 | 20,712 | 203 |
| K1Z | 39 | 22,714 | 172 |
| K2A | 31 | 17,156 | 181 |
| K2B | 25 | 33,254 | 75 |
| K2C | 29 | 29,359 | 99 |
| K2E | 12 | 20,172 | 59 |
| K2G | 76 | 50,692 | 150 |
| K2H | 32 | 26,759 | 120 |
| K2J | 74 | 81,863 | 90 |
| K2K | 34 | 24,922 | 136 |
| K2L | 38 | 18,739 | 203 |
| K2M | 12 | 28,708 | 42 |
| K2P | 68 | 18,825 | 361 |
| K2R | 0 | 1,237 | - |
| K2S | 63 | 38,903 | 162 |
| K2T | 29 | 11,871 | 244 |
| K2V | 8 | 10,486 | 76 |
| K2W | 3 | 8,553 | 35 |
| K4A | 41 | 62,125 | 66 |
| K4B | 2 | 4,704 | 43 |
| K4C | 0 | 4,538 | - |
| K4K | 17 | 16,958 | 100 |
| K4M | 22 | 14,380 | 153 |
| K4P | 8 | 10,448 | 77 |
| K4R | 3 | 8,740 | 34 |
| K6A | 15 | 12,529 | 120 |
| K6J | 18 | 17,827 | 101 |
| K6K | 0 | 3,452 | - |
| K6T | 0 | 690 | - |
| K6V | 35 | 28,174 | 124 |
| K7A | 19 | 17,575 | 108 |
| K7C | 29 | 20,382 | 142 |
| K7G | 8 | 8,740 | 92 |
| K7H | 13 | 16,175 | 80 |
| K7K | 26 | 33,705 | 77 |
| K7L | 31 | 19,621 | 158 |
| K7M | 48 | 49,268 | 97 |
| K7N | 4 | 8,395 | 48 |
| K7P | 43 | 24,660 | 174 |
| K7R | 21 | 15,480 | 136 |
| K7S | 14 | 14,265 | 98 |
| K7V | 18 | 13,943 | 129 |
| K8A | 44 | 26,392 | 167 |
| K8B | 0 | 490 | - |
| K8H | 12 | 16,188 | 74 |
| K8N | 55 | 30,393 | 181 |
| K8P | 24 | 23,166 | 104 |
| K8R | 0 | 2,012 | - |
| K8V | 32 | 28,936 | 111 |
| K9A | 40 | 26,757 | 149 |
| K9H | 43 | 29,704 | 145 |
| K9J | 73 | 44,547 | 164 |
| K9K | 9 | 13,359 | 67 |
| K9L | 1 | 7,813 | 13 |
| K9V | 29 | 29,369 | 99 |
| L0A | 7 | 15,325 | 46 |
| L0B | 3 | 15,501 | 19 |
| L0C | 2 | 9,486 | 21 |
| L0E | 8 | 21,898 | 37 |
| L0G | 39 | 44,945 | 87 |
| L0H | 0 | 1,223 | - |
| L0J | 8 | 4,945 | 162 |
| L0K | 14 | 38,386 | 36 |
| L0L | 18 | 42,022 | 43 |
| L0M | 27 | 37,226 | 73 |
| L0N | 0 | 2,395 | - |
| L0P | 2 | 7,779 | 26 |
| L0R | 61 | 83,602 | 73 |
| L0S | 62 | 60,409 | 103 |
| L1A | 18 | 15,845 | 114 |
| L1B | 20 | 13,203 | 151 |
| L1C | 48 | 50,217 | 96 |
| L1E | 26 | 28,325 | 92 |
| L1G | 49 | 43,497 | 113 |
| L1H | 60 | 32,298 | 186 |
| L1J | 44 | 43,107 | 102 |
| L1K | 33 | 43,574 | 76 |
| L1L | 5 | 12,976 | 39 |
| L1M | 27 | 22,542 | 120 |
| L1N | 77 | 49,649 | 155 |

| FSA | Dental Hygienists | Population | Dental Hygienists per 100,000 Population | FSA | Dental Hygienists | Population | Dental Hygienists per 100,000 Population |
|---|---|---|---|---|---|---|---|
| K6H | 29 | 30,440 | 95 | L1P | 15 | 22,928 | 65 |
| L1R | 44 | 42,221 | 104 | L5E | 2 | 12,972 | 15 |
| L1S | 45 | 42,045 | 107 | L5G | 34 | 20,714 | 164 |
| L1T | 48 | 52,632 | 91 | L5H | 23 | 17,409 | 132 |
| L1V | 58 | 52,787 | 110 | L5J | 21 | 28,208 | 74 |
| L1W | 19 | 18,433 | 103 | L5K | 12 | 13,363 | 90 |
| L1X | 10 | 24,639 | 41 | L5L | 42 | 43,585 | 96 |
| L1Y | 0 | 1,976 | - | L5M | 104 | 106,468 | 98 |
| L1Z | 28 | 31,989 | 88 | L5N | 73 | 83,700 | 87 |
| L2A | 15 | 17,293 | 87 | L5R | 41 | 37,022 | 111 |
| L2E | 23 | 18,587 | 124 | L5S | 5 | 27 | 18,519 |
| L2G | 32 | 30,344 | 105 | L5T | 1 | 19 | 5,263 |
| L2H | 24 | 29,820 | 80 | L5V | 42 | 49,149 | 85 |
| L2J | 25 | 14,908 | 168 | L5W | 31 | 24,145 | 128 |
| L2M | 17 | 33,704 | 50 | L6A | 91 | 89,287 | 102 |
| L2N | 58 | 30,719 | 189 | L6B | 14 | 34,763 | 40 |
| L2P | 4 | 15,128 | 26 | L6C | 29 | 51,738 | 56 |
| L2R | 33 | 25,558 | 129 | L6E | 17 | 38,387 | 44 |
| L2S | 26 | 19,400 | 134 | L6G | 2 | 8,364 | 24 |
| L2T | 19 | 11,401 | 167 | L6H | 82 | 72,662 | 113 |
| L2V | 40 | 19,795 | 202 | L6J | 69 | 24,420 | 283 |
| L2W | 2 | 1,959 | 102 | L6K | 20 | 14,118 | 142 |
| L3B | 14 | 25,869 | 54 | L6L | 29 | 30,012 | 97 |
| L3C | 54 | 33,462 | 161 | L6M | 61 | 72,547 | 84 |
| L3K | 10 | 19,458 | 51 | L6P | 33 | 91,155 | 36 |
| L3L | 0 | 879 | - | L6R | 55 | 93,783 | 59 |
| L3M | 39 | 28,433 | 137 | L6S | 38 | 54,361 | 70 |
| L3P | 44 | 37,336 | 118 | L6T | 61 | 39,614 | 154 |
| L3R | 116 | 62,060 | 187 | L6V | 22 | 42,392 | 52 |
| L3S | 26 | 55,403 | 47 | L6W | 45 | 23,547 | 191 |
| L3T | 35 | 50,405 | 69 | L6X | 53 | 76,113 | 70 |
| L3V | 53 | 48,651 | 109 | L6Y | 50 | 97,601 | 51 |
| L3X | 27 | 45,284 | 60 | L6Z | 40 | 33,888 | 118 |
| L3Y | 115 | 43,556 | 264 | L7A | 53 | 104,009 | 51 |
| L3Z | 38 | 41,072 | 93 | L7B | 11 | 17,809 | 62 |
| L4A | 40 | 50,389 | 79 | L7C | 12 | 33,846 | 35 |
| L4B | 36 | 36,080 | 100 | L7E | 35 | 34,516 | 101 |
| L4C | 124 | 74,636 | 166 | L7G | 51 | 48,051 | 106 |
| L4E | 37 | 54,606 | 68 | L7J | 7 | 13,766 | 51 |
| L4G | 74 | 62,061 | 119 | L7K | 2 | 8,486 | 24 |
| L4H | 68 | 77,353 | 88 | L7L | 53 | 47,433 | 112 |
| L4J | 92 | 75,120 | 122 | L7M | 63 | 48,498 | 130 |
| L4K | 49 | 20,172 | 243 | L7N | 39 | 13,160 | 296 |
| L4L | 94 | 55,426 | 170 | L7P | 25 | 29,777 | 84 |
| L4M | 86 | 46,902 | 183 | L7R | 40 | 17,338 | 231 |
| L4N | 137 | 100,835 | 136 | L7S | 9 | 12,207 | 74 |
| L4P | 27 | 29,861 | 90 | L7T | 18 | 18,555 | 97 |
| L4R | 36 | 19,245 | 187 | L8B | 18 | 25,778 | 70 |
| L4S | 25 | 36,688 | 68 | L8E | 30 | 41,927 | 72 |
| L4T | 20 | 38,182 | 52 | L8G | 66 | 21,805 | 303 |
| L4V | 1 | 5 | 20,000 | L8H | 11 | 26,187 | 42 |
| L4W | 29 | 19,744 | 147 | L8J | 20 | 29,146 | 69 |
| L4X | 10 | 18,719 | 53 | L8K | 21 | 32,598 | 64 |
| L4Y | 42 | 23,490 | 179 | L8L | 5 | 32,312 | 15 |
| L4Z | 44 | 37,218 | 118 | L8M | 20 | 13,774 | 145 |
| L5A | 43 | 47,406 | 91 | L8N | 19 | 15,568 | 122 |
| L5B | 102 | 67,131 | 152 | L8P | 52 | 24,773 | 210 |

| FSA | Dental Hygienists | Population | Dental Hygienists per 100,000 Population | FSA | Dental Hygienists | Population | Dental Hygienists per 100,000 Population |
|---|---|---|---|---|---|---|---|
| L5C | 27 | 29,284 | 92 | L8R | 10 | 11,651 | 86 |
| L8S | 12 | 15,787 | 76 | M3N | 22 | 40,846 | 54 |
| L8T | 12 | 19,405 | 62 | M4A | 9 | 14,589 | 62 |
| L8V | 25 | 21,165 | 118 | M4B | 9 | 18,612 | 48 |
| L8W | 19 | 26,077 | 73 | M4C | 25 | 46,440 | 54 |
| L9A | 52 | 25,899 | 201 | M4E | 23 | 25,473 | 90 |
| L9B | 29 | 25,025 | 116 | M4G | 26 | 19,598 | 133 |
| L9C | 44 | 41,202 | 107 | M4H | 13 | 18,698 | 70 |
| L9E | 2 | 16,933 | 12 | M4J | 18 | 35,128 | 51 |
| L9G | 52 | 25,020 | 208 | M4K | 39 | 30,913 | 126 |
| L9H | 28 | 31,221 | 90 | M4L | 14 | 32,218 | 43 |
| L9J | 4 | 2,272 | 176 | M4M | 26 | 25,052 | 104 |
| L9K | 22 | 15,928 | 138 | M4N | 19 | 16,058 | 118 |
| L9L | 21 | 14,620 | 144 | M4P | 53 | 25,057 | 212 |
| L9M | 4 | 16,607 | 24 | M4R | 7 | 11,909 | 59 |
| L9N | 14 | 21,656 | 65 | M4S | 65 | 30,754 | 211 |
| L9P | 22 | 17,383 | 127 | M4T | 45 | 10,332 | 436 |
| L9R | 36 | 25,949 | 139 | M4V | 21 | 19,273 | 109 |
| L9S | 26 | 33,434 | 78 | M4W | 37 | 15,296 | 242 |
| L9T | 117 | 110,956 | 105 | M4X | 12 | 19,896 | 60 |
| L9V | 9 | 17,750 | 51 | M4Y | 11 | 36,319 | 30 |
| L9W | 60 | 48,790 | 123 | M5A | 37 | 48,978 | 76 |
| L9X | 8 | 12,896 | 62 | M5B | 25 | 17,422 | 143 |
| L9Y | 41 | 28,925 | 142 | M5C | 13 | 3,149 | 413 |
| L9Z | 23 | 24,849 | 93 | M5E | 12 | 11,779 | 102 |
| M1B | 53 | 65,555 | 81 | M5G | 70 | 9,751 | 718 |
| M1C | 14 | 35,642 | 39 | M5H | 24 | 3,248 | 739 |
| M1E | 19 | 48,033 | 40 | M5J | 29 | 16,879 | 172 |
| M1G | 3 | 30,894 | 10 | M5M | 26 | 25,495 | 102 |
| M1H | 51 | 23,964 | 213 | M5N | 11 | 16,154 | 68 |
| M1J | 15 | 37,002 | 41 | M5P | 11 | 19,791 | 56 |
| M1K | 31 | 48,175 | 64 | M5R | 60 | 26,197 | 229 |
| M1L | 17 | 35,833 | 47 | M5S | 29 | 17,074 | 170 |
| M1M | 13 | 23,258 | 56 | M5T | 27 | 17,903 | 151 |
| M1N | 18 | 22,976 | 78 | M5V | 54 | 59,912 | 90 |
| M1P | 53 | 45,170 | 117 | M6A | 36 | 22,380 | 161 |
| M1R | 40 | 30,467 | 131 | M6B | 25 | 28,534 | 88 |
| M1S | 37 | 37,663 | 98 | M6C | 22 | 23,997 | 92 |
| M1T | 33 | 34,607 | 95 | M6E | 22 | 37,684 | 58 |
| M1V | 27 | 50,825 | 53 | M6G | 27 | 30,600 | 88 |
| M1W | 44 | 47,070 | 93 | M6H | 25 | 43,730 | 57 |
| M1X | 7 | 14,810 | 47 | M6J | 19 | 32,111 | 59 |
| M2H | 13 | 23,396 | 56 | M6K | 23 | 42,110 | 55 |
| M2J | 53 | 61,761 | 86 | M6L | 6 | 20,156 | 30 |
| M2K | 51 | 25,377 | 201 | M6M | 24 | 42,148 | 57 |
| M2L | 9 | 11,274 | 80 | M6N | 17 | 40,252 | 42 |
| M2M | 16 | 31,915 | 50 | M6P | 30 | 39,870 | 75 |
| M2N | 106 | 75,100 | 141 | M6R | 12 | 19,349 | 62 |
| M2P | 16 | 7,546 | 212 | M6S | 62 | 34,029 | 182 |
| M2R | 22 | 40,581 | 54 | M7A | 1 | 5 | 20,000 |
| M3A | 9 | 34,361 | 26 | M8V | 26 | 44,144 | 59 |
| M3B | 7 | 12,880 | 54 | M8W | 17 | 22,381 | 76 |
| M3C | 31 | 39,616 | 78 | M8X | 43 | 10,624 | 405 |
| M3H | 32 | 38,416 | 83 | M8Y | 25 | 21,986 | 114 |
| M3J | 61 | 26,600 | 229 | M8Z | 26 | 19,312 | 135 |
| M3K | 19 | 7,865 | 242 | M9A | 18 | 36,642 | 49 |
| M3L | 11 | 19,263 | 57 | M9B | 36 | 33,236 | 108 |

| FSA | Dental Hygienists | Population | Dental Hygienists per 100,000 Population | FSA | Dental Hygienists | Population | Dental Hygienists per 100,000 Population |
|---|---|---|---|---|---|---|---|
| M3M | 33 | 24,906 | 132 | M9C | 21 | 38,725 | 54 |
| M9L | 8 | 11,737 | 68 | N3T | 24 | 37,406 | 64 |
| M9M | 6 | 24,245 | 25 | N3V | 0 | 1,562 | - |
| M9N | 13 | 26,059 | 50 | N3W | 16 | 15,805 | 101 |
| M9P | 31 | 20,645 | 150 | N3Y | 33 | 23,402 | 141 |
| M9R | 14 | 33,783 | 41 | N4B | 2 | 8,223 | 24 |
| M9V | 36 | 53,878 | 67 | N4G | 24 | 21,519 | 112 |
| M9W | 30 | 40,977 | 73 | N4K | 37 | 28,975 | 128 |
| N0A | 26 | 31,957 | 81 | N4L | 4 | 8,337 | 48 |
| N0B | 36 | 86,919 | 41 | N4N | 23 | 10,341 | 222 |
| N0C | 5 | 17,537 | 29 | N4S | 41 | 33,055 | 124 |
| N0E | 11 | 36,557 | 30 | N4T | 8 | 15,161 | 53 |
| N0G | 46 | 86,935 | 53 | N4V | 0 | 3,477 | - |
| N0H | 41 | 53,674 | 76 | N4W | 10 | 13,311 | 75 |
| N0J | 9 | 33,457 | 27 | N4X | 6 | 9,946 | 60 |
| N0K | 15 | 24,817 | 60 | N4Z | 0 | 2,362 | - |
| N0L | 31 | 49,817 | 62 | N5A | 37 | 32,898 | 112 |
| N0M | 49 | 69,177 | 71 | N5C | 10 | 15,721 | 64 |
| N0N | 22 | 42,297 | 52 | N5H | 8 | 15,625 | 51 |
| N0P | 24 | 53,140 | 45 | N5L | 1 | 3,714 | 27 |
| N0R | 25 | 50,203 | 50 | N5P | 19 | 21,081 | 90 |
| N1A | 9 | 12,614 | 71 | N5R | 21 | 30,680 | 68 |
| N1C | 0 | 3,678 | - | N5V | 14 | 32,961 | 42 |
| N1E | 25 | 44,799 | 56 | N5W | 21 | 24,740 | 85 |
| N1G | 57 | 29,688 | 192 | N5X | 29 | 36,963 | 78 |
| N1H | 51 | 42,178 | 121 | N5Y | 51 | 35,934 | 142 |
| N1K | 3 | 10,908 | 28 | N5Z | 15 | 24,067 | 62 |
| N1L | 19 | 17,474 | 109 | N6A | 53 | 13,810 | 384 |
| N1M | 19 | 18,498 | 103 | N6B | 28 | 11,101 | 252 |
| N1P | 0 | 7,991 | - | N6C | 21 | 30,629 | 69 |
| N1R | 59 | 42,928 | 137 | N6E | 29 | 28,140 | 103 |
| N1S | 22 | 20,171 | 109 | N6G | 37 | 50,940 | 73 |
| N1T | 6 | 18,513 | 32 | N6H | 63 | 44,215 | 142 |
| N2A | 32 | 32,454 | 99 | N6J | 29 | 27,497 | 105 |
| N2B | 11 | 16,939 | 65 | N6K | 55 | 37,596 | 146 |
| N2C | 24 | 17,681 | 136 | N6L | 1 | 5,167 | 19 |
| N2E | 14 | 40,428 | 35 | N6M | 4 | 8,542 | 47 |
| N2G | 48 | 14,580 | 329 | N6N | 2 | 728 | 275 |
| N2H | 22 | 22,456 | 98 | N6P | 13 | 10,943 | 119 |
| N2J | 59 | 20,899 | 282 | N7A | 14 | 12,495 | 112 |
| N2K | 26 | 29,381 | 88 | N7G | 22 | 19,921 | 110 |
| N2L | 31 | 37,953 | 82 | N7L | 36 | 25,127 | 143 |
| N2M | 32 | 36,494 | 88 | N7M | 18 | 25,579 | 70 |
| N2N | 28 | 26,596 | 105 | N7S | 39 | 28,410 | 137 |
| N2P | 20 | 25,040 | 80 | N7T | 33 | 26,033 | 127 |
| N2R | 7 | 18,445 | 38 | N7V | 19 | 11,611 | 164 |
| N2T | 19 | 20,634 | 92 | N7W | 0 | 2,578 | - |
| N2V | 4 | 19,428 | 21 | N7X | 0 | 1,129 | - |
| N2Z | 16 | 12,341 | 130 | N8A | 13 | 12,624 | 103 |
| N3A | 17 | 17,045 | 100 | N8H | 26 | 27,530 | 94 |
| N3B | 10 | 13,190 | 76 | N8M | 10 | 11,819 | 85 |
| N3C | 23 | 27,518 | 84 | N8N | 37 | 24,534 | 151 |
| N3E | 0 | 2,802 | - | N8P | 1 | 14,603 | 7 |
| N3H | 6 | 24,306 | 25 | N8R | 14 | 12,733 | 110 |
| N3L | 19 | 17,676 | 107 | N8S | 11 | 22,354 | 49 |
| N3P | 0 | 8,542 | - | N8T | 29 | 17,935 | 162 |
| N3R | 52 | 35,858 | 145 | N8W | 45 | 24,057 | 187 |

| FSA | Dental Hygienists | Population | Dental Hygienists per 100,000 Population | FSA | Dental Hygienists | Population | Dental Hygienists per 100,000 Population |
|---|---|---|---|---|---|---|---|
| N3S | 24 | 27,527 | 87 | N8X | 49 | 16,457 | 298 |
| N8Y | 15 | 19,254 | 78 | P1P | 8 | 11,083 | 72 |
| N9A | 35 | 26,107 | 134 | P2A | 23 | 14,695 | 157 |
| N9B | 8 | 19,613 | 41 | P2B | 16 | 9,754 | 164 |
| N9C | 9 | 12,538 | 72 | P2N | 4 | 6,944 | 58 |
| N9E | 15 | 21,465 | 70 | P3A | 64 | 24,326 | 263 |
| N9G | 22 | 20,712 | 106 | P3B | 4 | 16,434 | 24 |
| N9H | 23 | 13,173 | 175 | P3C | 20 | 17,596 | 114 |
| N9J | 15 | 20,089 | 75 | P3E | 60 | 29,132 | 206 |
| N9K | 3 | 2,757 | 109 | P3G | 0 | 4,167 | - |
| N9V | 13 | 21,688 | 60 | P3L | 0 | 7,996 | - |
| N9Y | 17 | 15,836 | 107 | P3N | 13 | 7,980 | 163 |
| P0A | 6 | 17,822 | 34 | P3P | 10 | 15,408 | 65 |
| P0B | 2 | 9,589 | 21 | P3Y | 4 | 7,757 | 52 |
| P0C | 1 | 4,824 | 21 | P4N | 26 | 24,098 | 108 |
| P0E | 2 | 3,606 | 55 | P4P | 2 | 3,232 | 62 |
| P0G | 0 | 3,553 | - | P4R | 6 | 4,795 | 125 |
| P0H | 15 | 31,909 | 47 | P5A | 7 | 11,372 | 62 |
| P0J | 23 | 20,179 | 114 | P5E | 8 | 5,427 | 147 |
| P0K | 5 | 11,456 | 44 | P5N | 11 | 8,260 | 133 |
| P0L | 23 | 22,859 | 101 | P6A | 61 | 33,800 | 180 |
| P0M | 25 | 49,727 | 50 | P6B | 36 | 22,754 | 158 |
| P0N | 5 | 9,435 | 53 | P6C | 3 | 18,766 | 16 |
| P0P | 10 | 20,285 | 49 | P7A | 24 | 28,587 | 84 |
| P0R | 7 | 12,976 | 54 | P7B | 67 | 22,675 | 295 |
| P0S | 4 | 10,170 | 39 | P7C | 34 | 21,695 | 157 |
| P0T | 18 | 32,000 | 56 | P7E | 12 | 20,897 | 57 |
| P0V | 3 | 22,944 | 13 | P7G | 2 | 13,238 | 15 |
| P0W | 1 | 6,454 | 15 | P7J | 0 | 4,859 | - |
| P0X | 2 | 8,410 | 24 | P7K | 4 | 6,801 | 59 |
| P0Y | 0 | 96 | - | P7L | 0 | 2,188 | - |
| P1A | 6 | 16,959 | 35 | P8N | 14 | 9,619 | 146 |
| P1B | 74 | 33,113 | 223 | P8T | 7 | 5,541 | 126 |
| P1C | 0 | 3,371 | - | P9A | 16 | 10,079 | 159 |
| P1H | 28 | 18,099 | 155 | P9N | 17 | 14,088 | 121 |
| P1L | 14 | 17,984 | 78 | | | | |

FSA – Forward Sortation Area

#### Additional File 5: Dental Hygienists, Population and Dental Hygienist Rate per 50 FSA-2s

| FSA-2 | Dental Hygienists | Population | Dental Hygienists per 100,000 Population |
|---|---|---|---|
| K0 | 295 | 594,410 | 50 |
| K1 | 540 | 444,717 | 121 |
| K2 | 534 | 421,499 | 127 |
| K3 | 0 | 0 | - |
| K4 | 93 | 121,893 | 76 |
| K5 | 0 | 0 | - |
| K6 | 97 | 93,112 | 104 |
| K7 | 274 | 242,209 | 113 |
| K8 | 167 | 127,577 | 131 |
| K9 | 195 | 151,549 | 129 |
| L0 | 251 | 385,142 | 65 |
| L1 | 674 | 644,883 | 105 |
| L2 | 318 | 268,616 | 118 |
| L3 | 571 | 491,868 | 116 |
| L4 | 1071 | 876,732 | 122 |
| L5 | 603 | 580,602 | 104 |
| L6 | 811 | 988,752 | 82 |
| L7 | 418 | 447,451 | 93 |
| L8 | 340 | 357,953 | 95 |
| L9 | 614 | 557,315 | 110 |
| M0 | 0 | 0 | - |
| M1 | 475 | 631,944 | 75 |
| M2 | 286 | 276,950 | 103 |
| M3 | 225 | 244,753 | 92 |
| M4 | 472 | 451,615 | 105 |
| M5 | 428 | 293,732 | 146 |
| M6 | 350 | 456,950 | 77 |
| M7 | 0 | 0 | - |
| M8 | 137 | 118,447 | 116 |
| M9 | 213 | 319,927 | 67 |
| N0 | 340 | 636,487 | 53 |
| N1 | 270 | 269,440 | 100 |
| N2 | 393 | 391,749 | 100 |
| N3 | 224 | 252,639 | 89 |
| N4 | 155 | 154,707 | 100 |
| N5 | 226 | 274,384 | 82 |
| N6 | 335 | 269,308 | 124 |
| N7 | 181 | 152,883 | 118 |
| N8 | 250 | 203,900 | 123 |
| N9 | 160 | 173,978 | 92 |
| P0 | 152 | 298,294 | 51 |
| P1 | 130 | 100,609 | 129 |
| P2 | 43 | 31,393 | 137 |
| P3 | 175 | 130,796 | 134 |
| P4 | 34 | 32,125 | 106 |
| P5 | 26 | 25,059 | 104 |
| P6 | 100 | 75,320 | 133 |
| P7 | 143 | 120,940 | 118 |
| P8 | 21 | 15,160 | 139 |
| P9 | 33 | 24,167 | 137 |

#### Additional File 6: Dental Hygienists, Population and Dental Hygienist Rate per 5 FSA-1s

| FSA-1 (Postal District) | Dental Hygienists | Population | Dental Hygienists per 100,000 Population |
|---|---|---|---|
| K | 2,195 | 2,196,966 | 100 |
| L | 5,671 | 5,599,314 | 101 |
| M | 2,586 | 2,794,318 | 93 |
| N | 2,534 | 2,779,475 | 91 |
| P | 857 | 853,863 | 100 |

#### Additional File 7: Dental Hygienists per Forward Sortation Area (FSA) in Ontario

##### Figure AF7a: Dental Hygienists per Forward Sortation Area (FSA) in Ontario

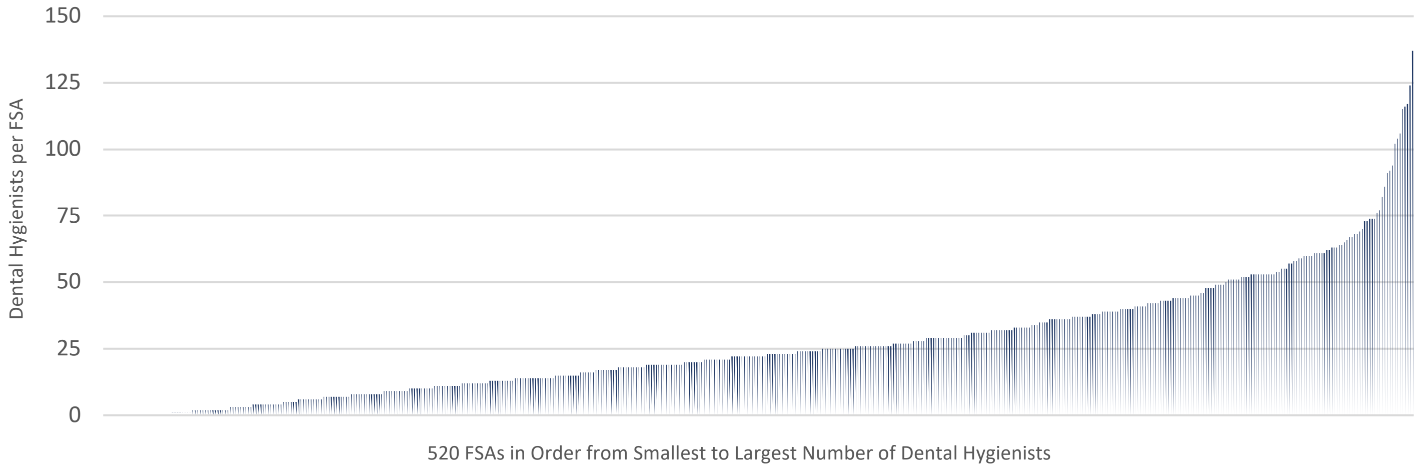


##### Figure AF7b: Dental Hygienists per Forward Sortation Area-2 (FSA-2) in Ontario

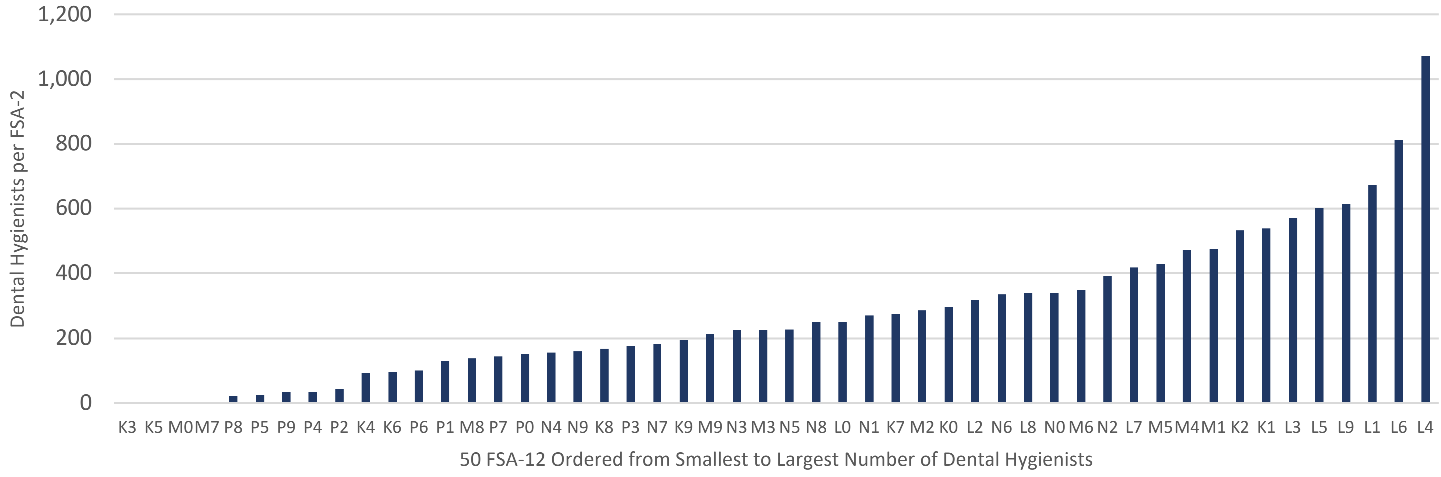


##### Figure AF7c: Dental Hygienists per Forward Sortation Area-1 (FSA-1) in Ontario

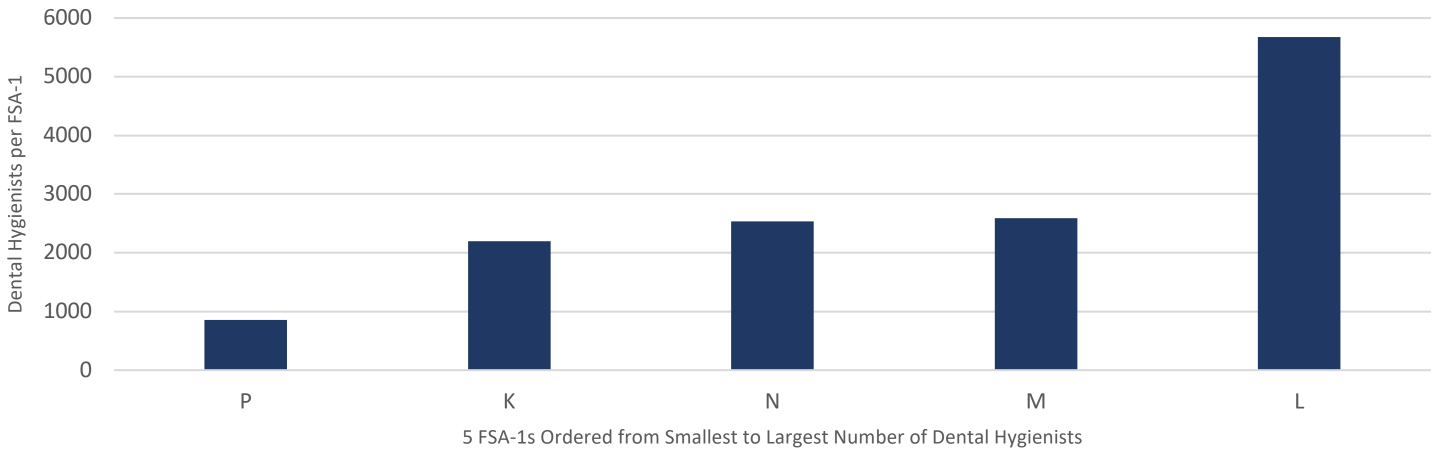

#### Additional File 8: Heat Maps

**Heat Maps 1a/1b** provide a comparative overview of the FSA and FSA-2 maps for the entire province of Ontario. Note that there are two distinct legend-scales for the FSA and FSA-2 maps. Yellow regions on the map indicate a dental hygienist rate of zero with progressively darker colours indicating higher dental hygienist rates, with purple regions having the highest dental hygienist rates overall.

**Heat Map 2** provides a larger version of Heat Map 1a to make additional FSAs in the province visible. However, many urban areas of Heat Map 2 are still too small to visualize well. Therefore, **Heat Maps 3-7** provide zoomed-in versions of the FSA maps for each of the five Ontario postal districts, allowing each individual FSA to be visualized geographically. Note that each of these maps also includes a small Ontario map with a red circle indicating which part of the province the zoomed-in map represents.

A few observations of the FSA level heat maps are provided below:

- The K district representing eastern Ontario (**Heat Map 3**), includes seven FSAs that have a zero dental hygienist rate, while its highest dental hygienist rates are in the Ottawa region.
- The L district representing central Ontario (**Heat Map 4**), includes four FSAs that have a zero dental hygienist rate, with some of these in urban areas and therefore requiring further investigation. L district's highest dental hygienist rates are all in urban areas, including in Hamilton, Oakville, Burlington, Newmarket and Concord. Mississauga, also in the L district, has a couple of the highest dental hygienist rates in the entire province, however, these FSAs are connected to Toronto Pearson International Airport, having very small populations whereby only a few dental hygienists in that FSA dramatically increase the dental hygienist rate.
- The M district representing metropolitan Toronto (**Heat Map 5**), has no FSAs with a zero dental hygienist rate, which is consistent with the overall results as the M district is the only one of the five that has no rural or remote areas. Unsurprisingly, the M district also contains some of the highest dental hygienist rates in Ontario given it is the most densely populated region in the province. However, as with the other districts, the M district still has some variation across FSAs in the region.
- The N district representing southwestern Ontario (**Heat Map 6**), includes nine FSAs with a zero dental hygienist rate. The N district's highest dental hygienist rate FSAs are mostly with in urban areas (e.g., London, Kitchener-Waterloo, Windsor), with one exception in a less urban area near Hanover, Ontario (this could be a target for further investigation to understand why the dental hygienist rate is higher in this FSA).
- The P district representing northern Ontario (**Heat Map 7**), includes seven FSAs with a zero dental hygienist rate. Northern Ontario covers a huge geography and is sparsely populated, so the heat map must be interpreted cautiously, however, the results are consistent with the rest of the province with the more rural/remote FSAs having lower dental hygienist rates and the higher dental hygienist rate FSAs connected to urban areas in Thunder Bay, Sudbury and North Bay.

#### Heat Map 1a: Dental Hygienists per 100,000 Population for All Ontario FSAs (Comparative Overview)

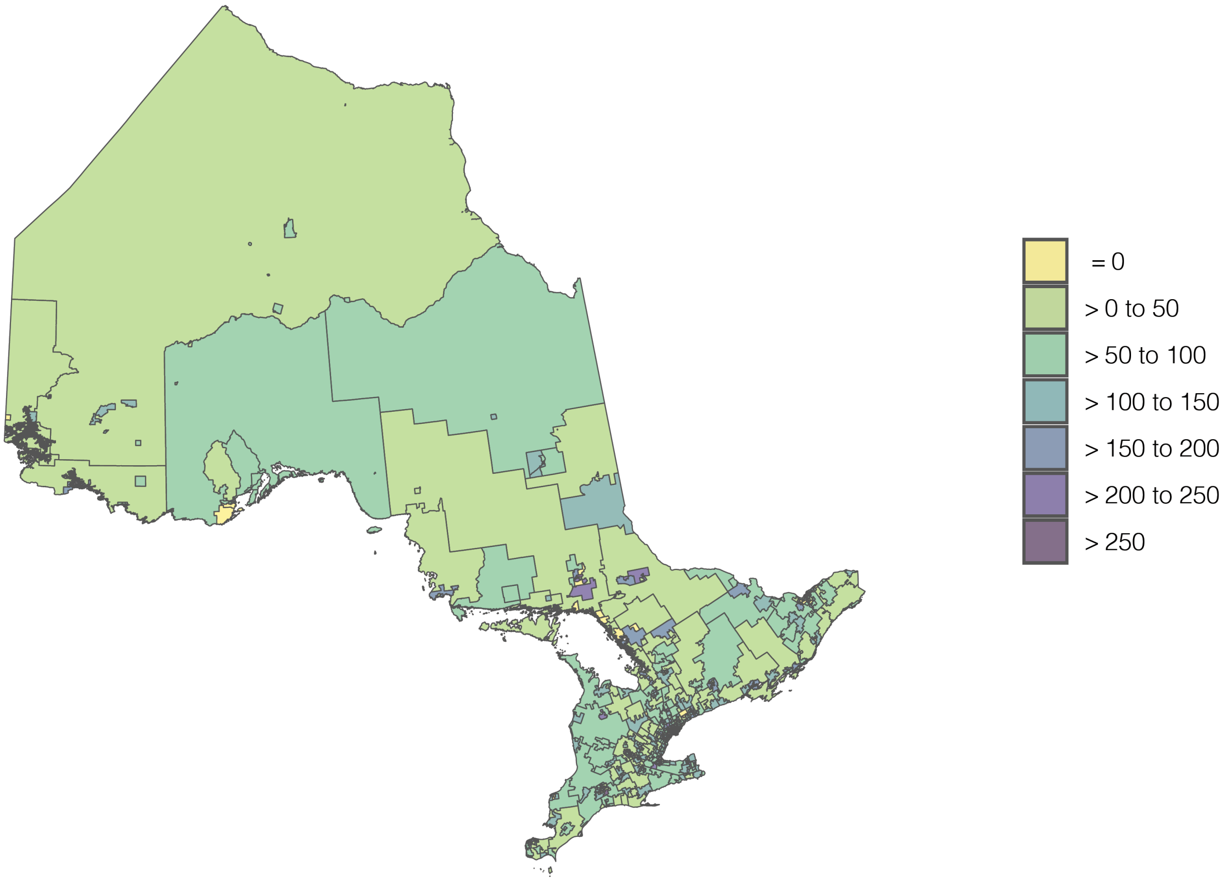


#### Heat Map 1b: Dental Hygienists per 100,000 Population for All Ontario FSA-2s (Comparative Overview)

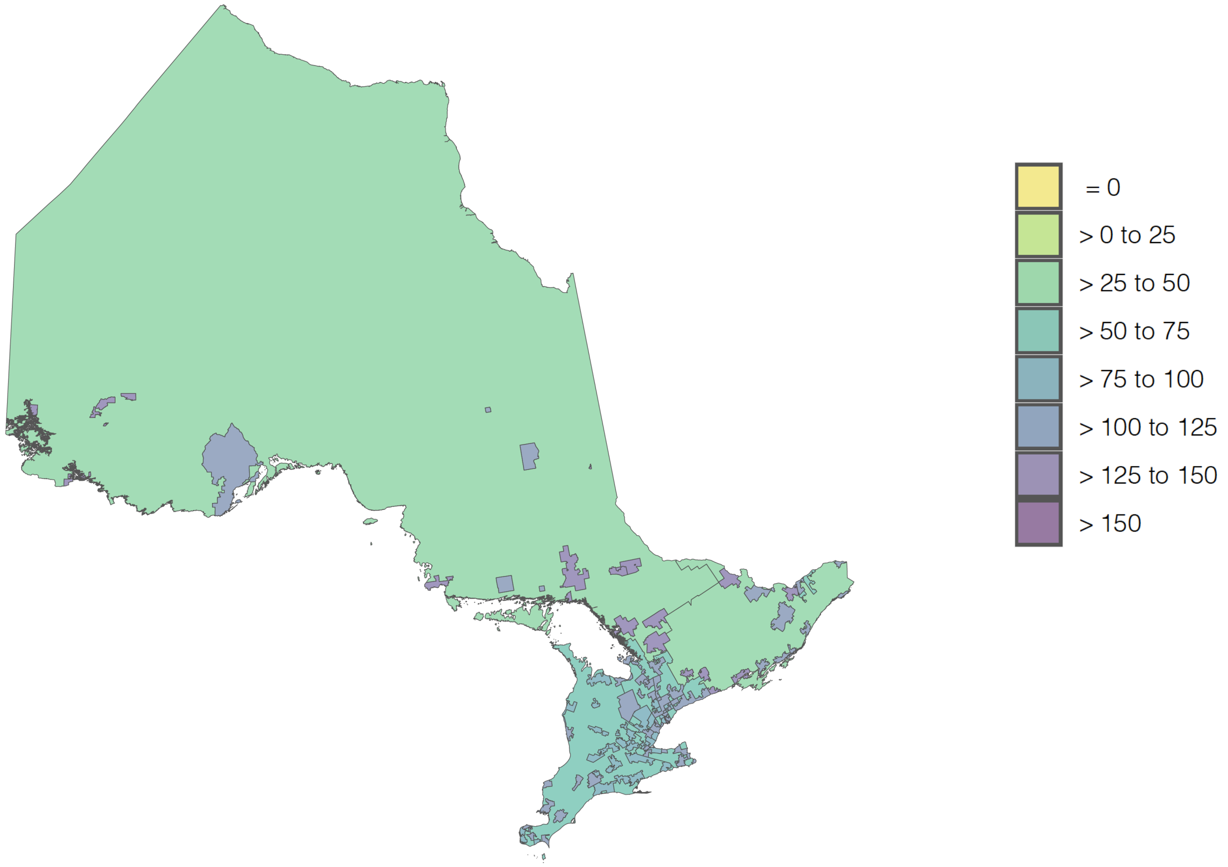

##### Heat Map 2: Dental Hygienists per 100,000 Population for All Ontario FSAs

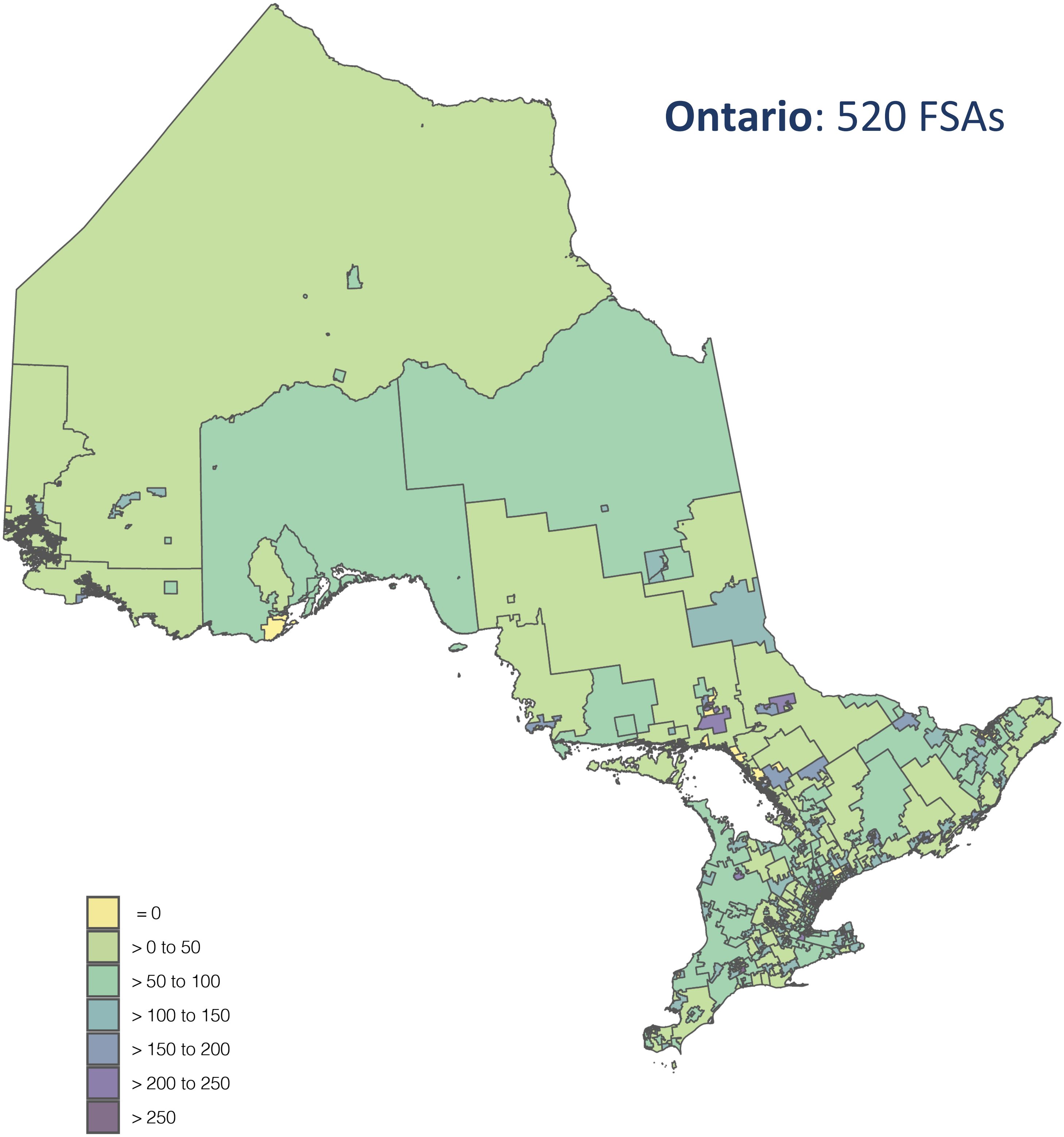

##### Heat Map 3: Dental Hygienists per 100,000 Population for K FSAs

#### **K**: 84 FSAs

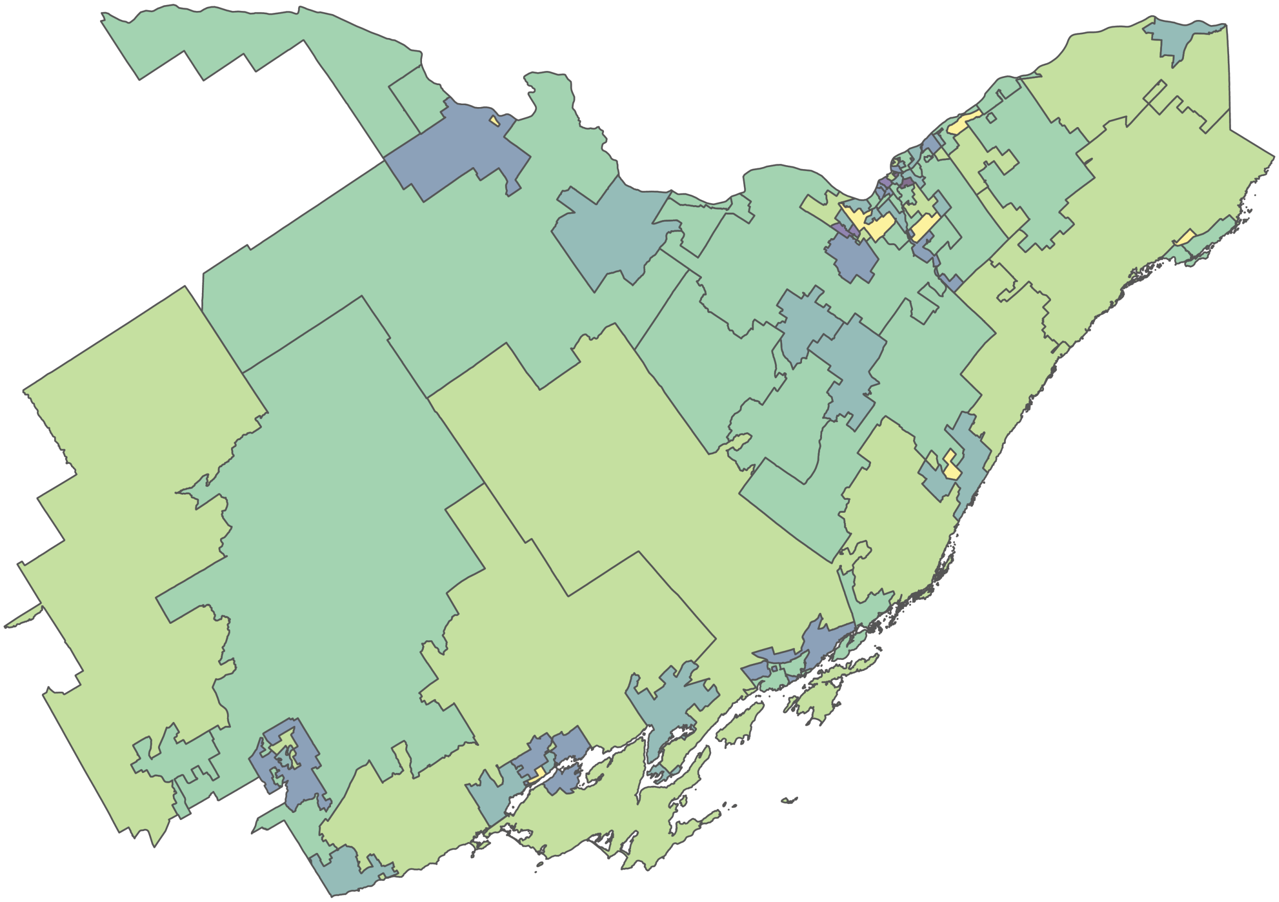

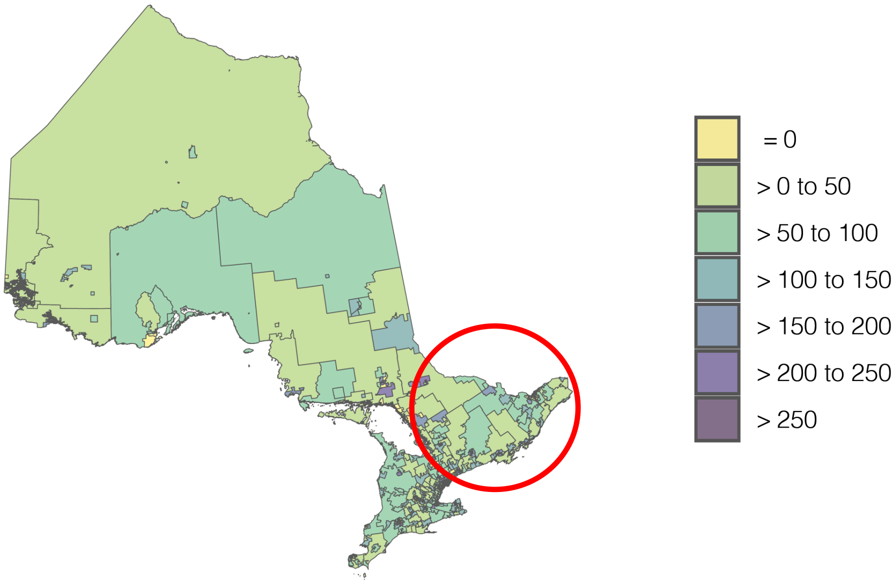

##### Heat Map 4: Dental Hygienists per 100,000 Population for L FSAs

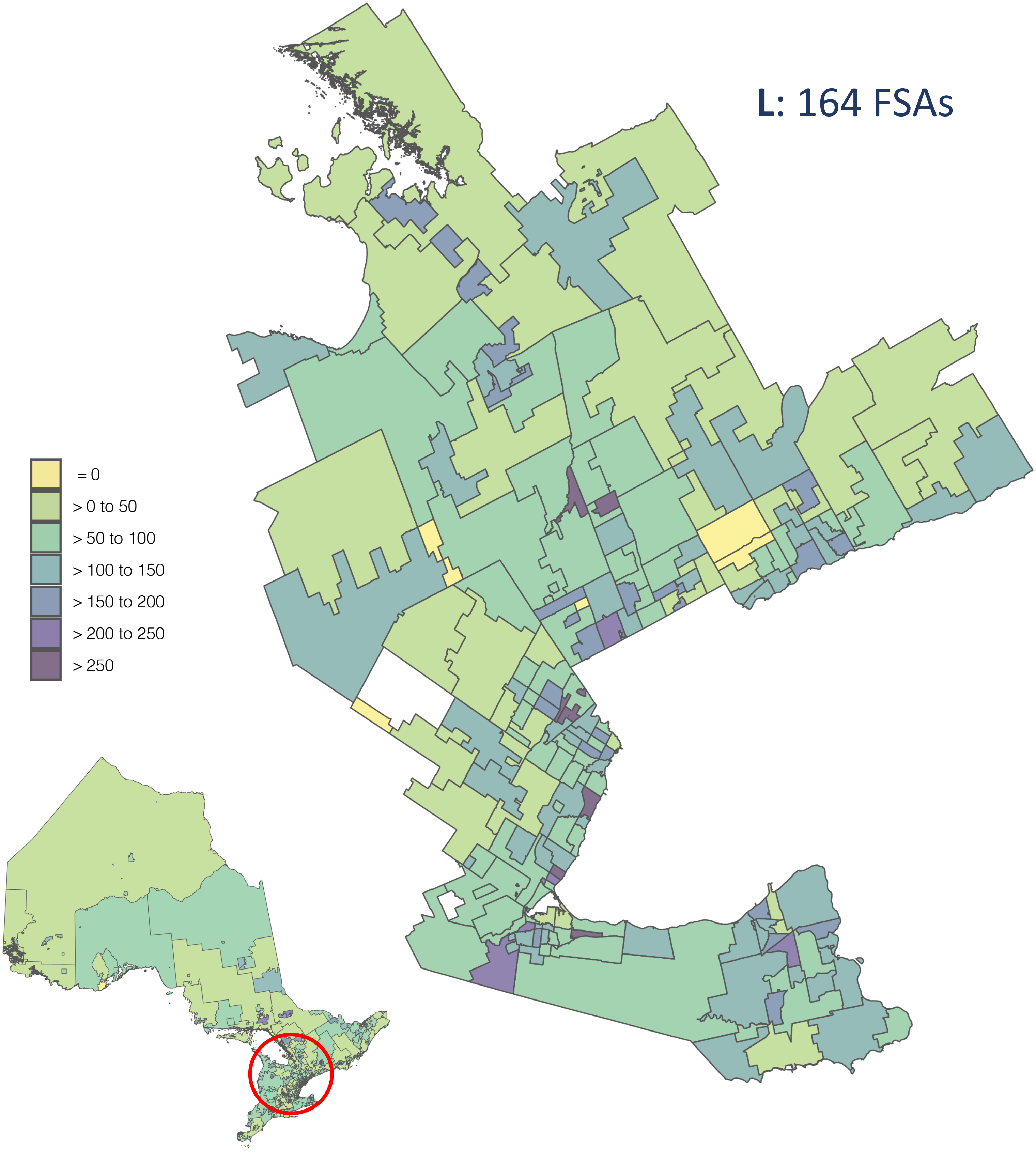

**Heat Map 5: Dental Hygienists per 100,000 Population for M FSAs**

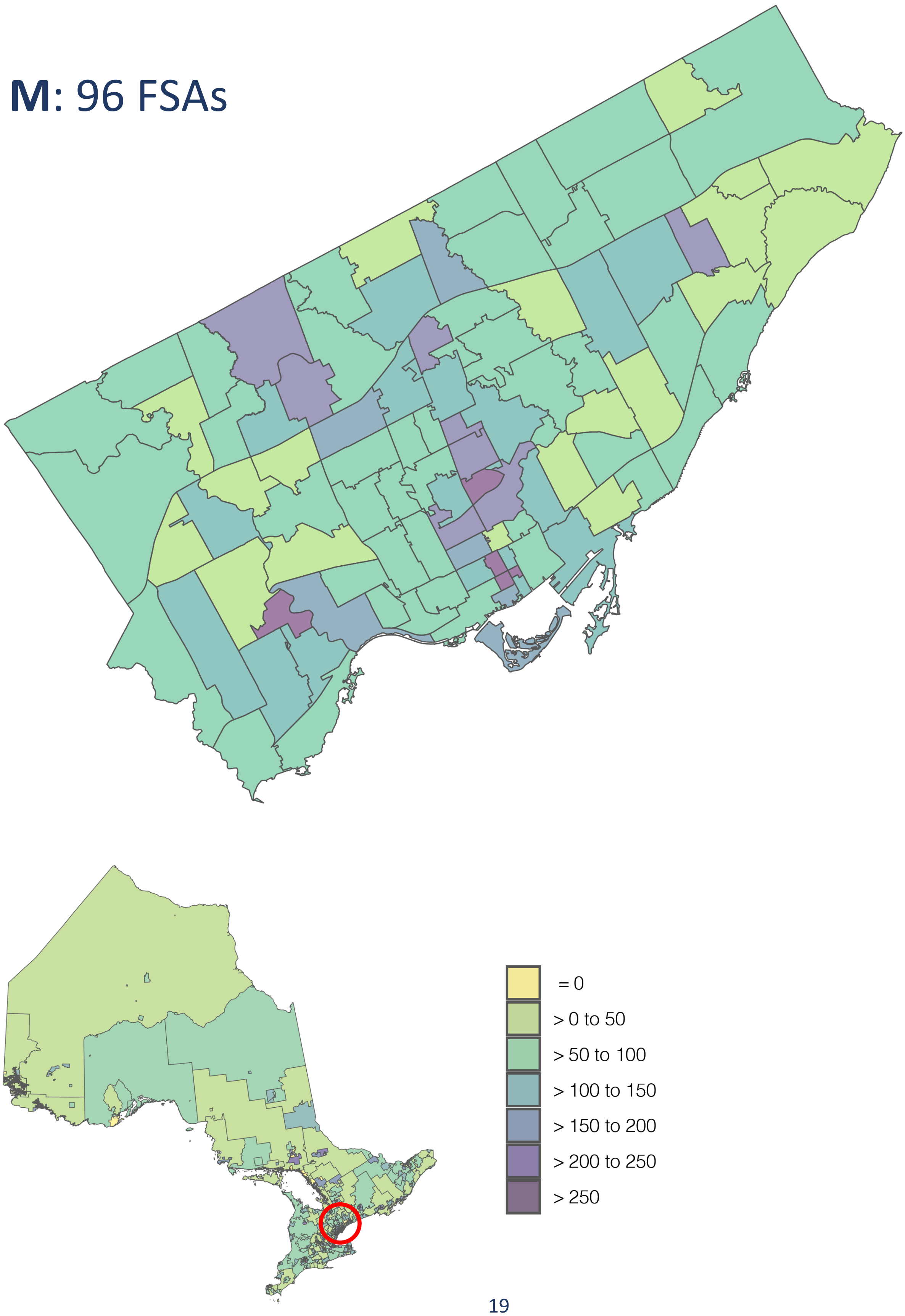

**Heat Map 6: Dental Hygienists per 100,000 Population for N FSAs**

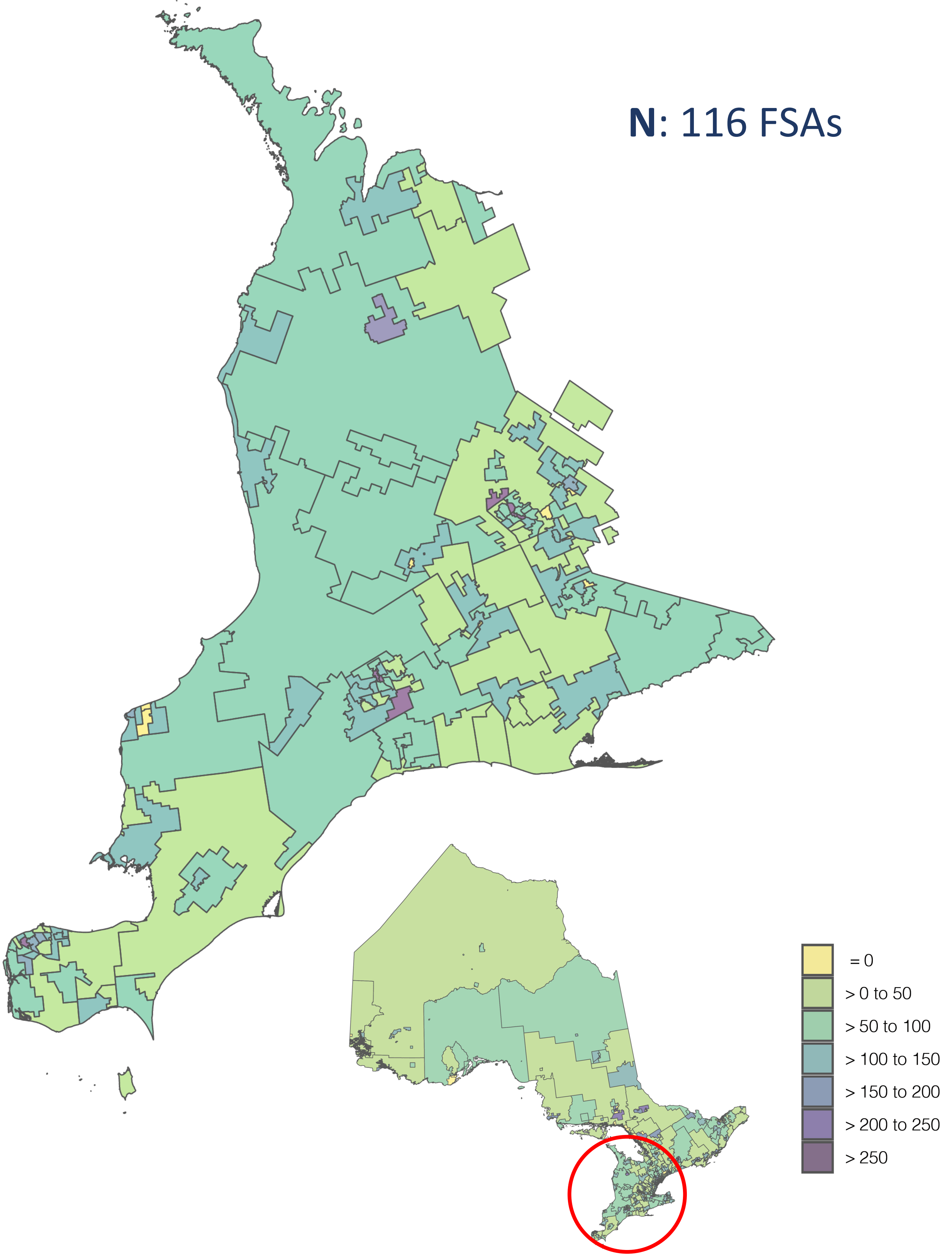

##### Heat Map 7: Dental Hygienists per 100,000 Population for P FSAs

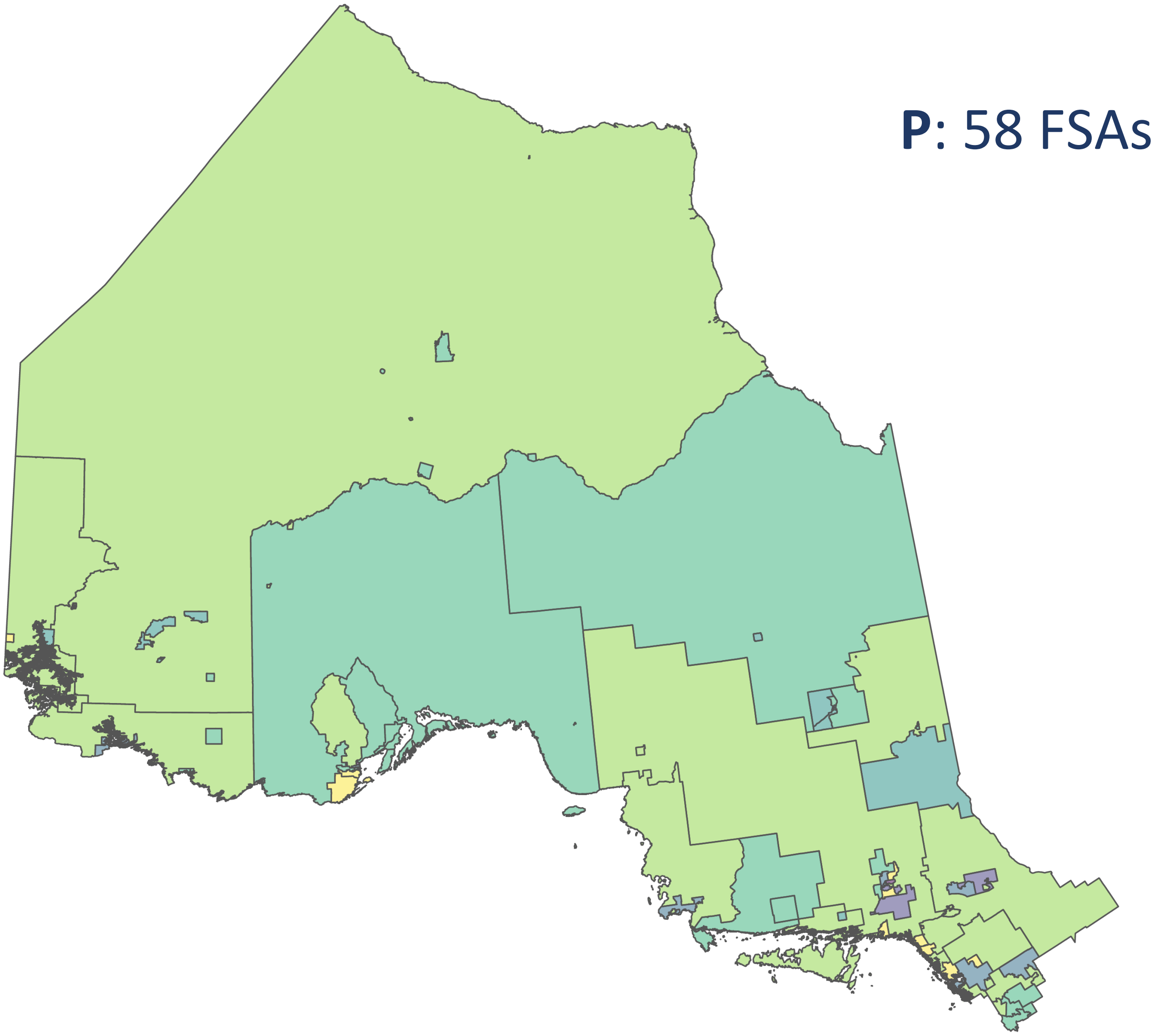


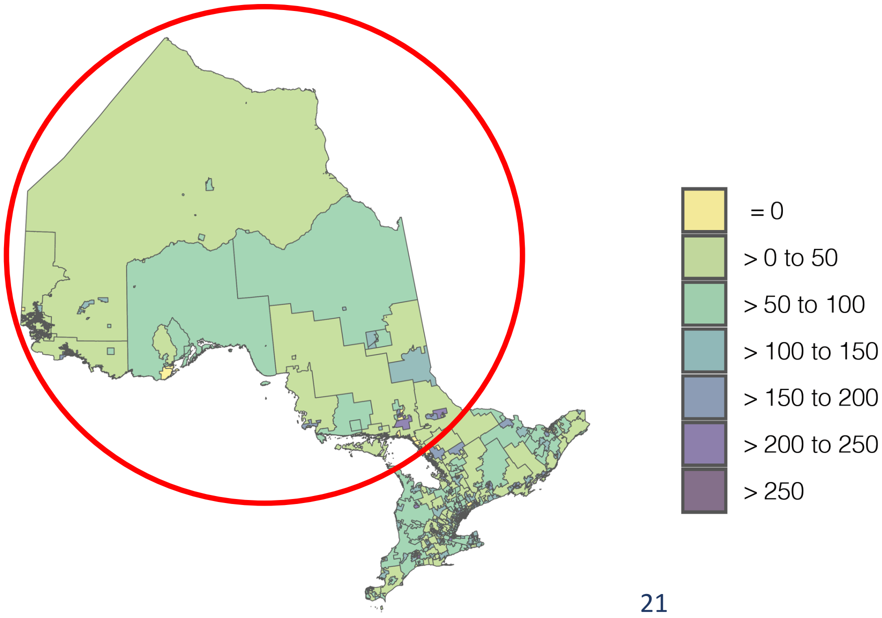

The second series of maps displays the same data at the more aggregated FSA-2 level of analysis, again for all of Ontario and then zoomed-in for each of the five postal districts in the province. **Heat Map 8** provides a larger version of Heat Map 1b to make additional FSA-2s in the province visible. **Heat Maps 9-13** provide zoomed-in versions of the FSA-2 maps for each of the five Ontario postal districts.

A few observations on the FSA-2 level heat maps are provided below:

- The K district (**Heat Map 9**) is dominated geographically by the 'K0' FSA-2, which has the lowest dental hygienist rate in the district (50 dental hygienists per 100,000 population). There are a few pockets of higher dental hygienist rates, aligned with urban areas in Ottawa, however four of the FSA-2s have multiple non-contiguous regions which makes the interpretation of the results more challenging.
- The L district (**Heat Map 10**) is less dominated geographically by the 'L0' FSA-2, which has a much higher dental hygienist rate for its rural/remote FSA-2 (65 dental hygienists per 100,000 population) than the 'K0' (50), 'N0' (53) and 'P0' (51) FSA-2s. This is likely because the L district has less rural/remote areas relative to the K, N or P districts. The L district has five FSA-2s that have multiple non-contiguous regions spread across the larger district, which makes interpretation of results more challenging, however, the heat maps do help to identify higher dental hygienist rates in the Mississauga and Hamilton urban areas of the district.
- The M district (**Heat Map 11**) does not have a rural/remote FSA-2, and among the eight FSA-2s in the dataset, all but one represents contiguous geographic areas in the metropolitan Toronto area. The core FSA-2 ('M5') that runs along a central north-south corridor from Lake Ontario to the 401 highway has the highest dental hygienist rate for an FSA-2 in the province (146 dental hygienists per 100,000 population).
- The N district (**Heat Map 12**) is dominated geographically by the 'N0' FSA-2, which has the lowest dental hygienist rate in the district (53 dental hygienists per 100,000 population). Most of the remaining FSA-2s capture multiple non-contiguous areas with only London and Windsor identifiable among the regions with higher dental hygienist rates.
- The P district (**Heat Map 13**) is dominated geographically by the 'P0' FSA-2, which has the lowest dental hygienist rate in the district (51 dental hygienist per 100,000 population). There are a few identifiable pockets with higher dental hygienist rates, including for Thunder Bay, Sudbury and Timmins, but several of P district's FSA-2s have multiple non-contiguous regions spread across the district, making interpretation of the results more challenging.

##### Heat Map 8: Dental Hygienists per 100,000 Population for All Ontario FSA-2s

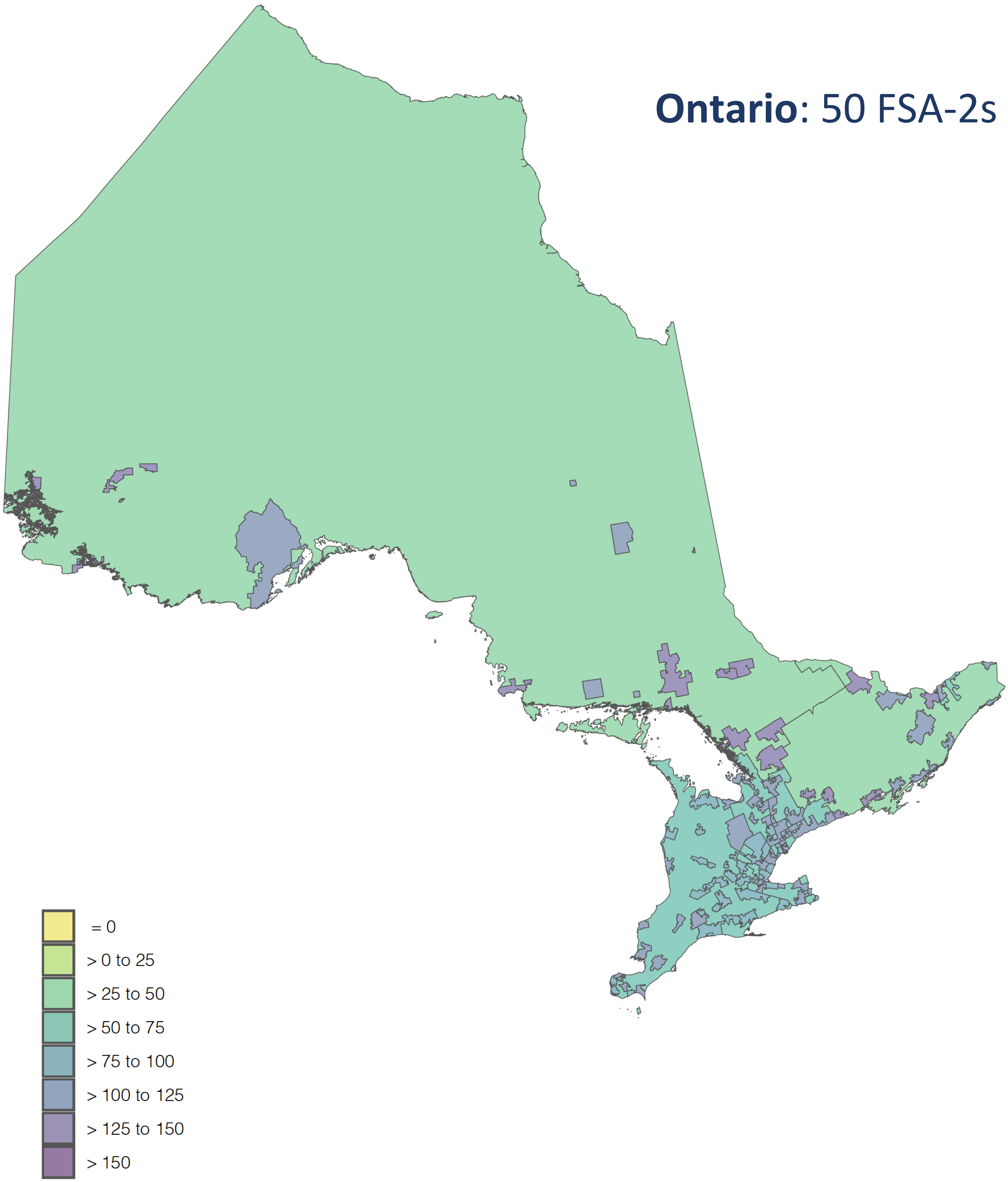

##### Heat Map 9: Dental Hygienists per 100,000 Population for K FSA-2s

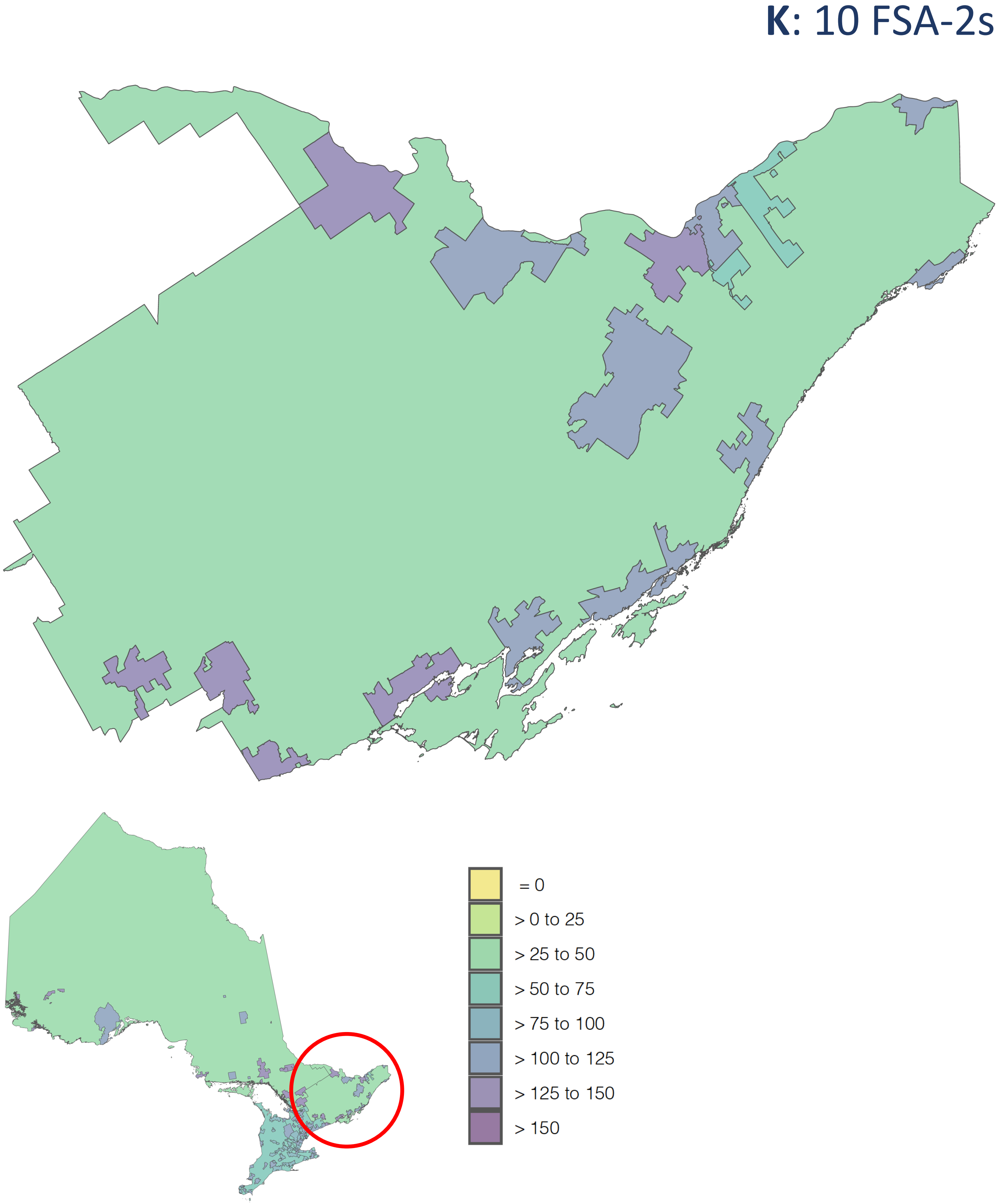

**Heat Map 10: Dental Hygienists per 100,000 Population for L FSA-2s**

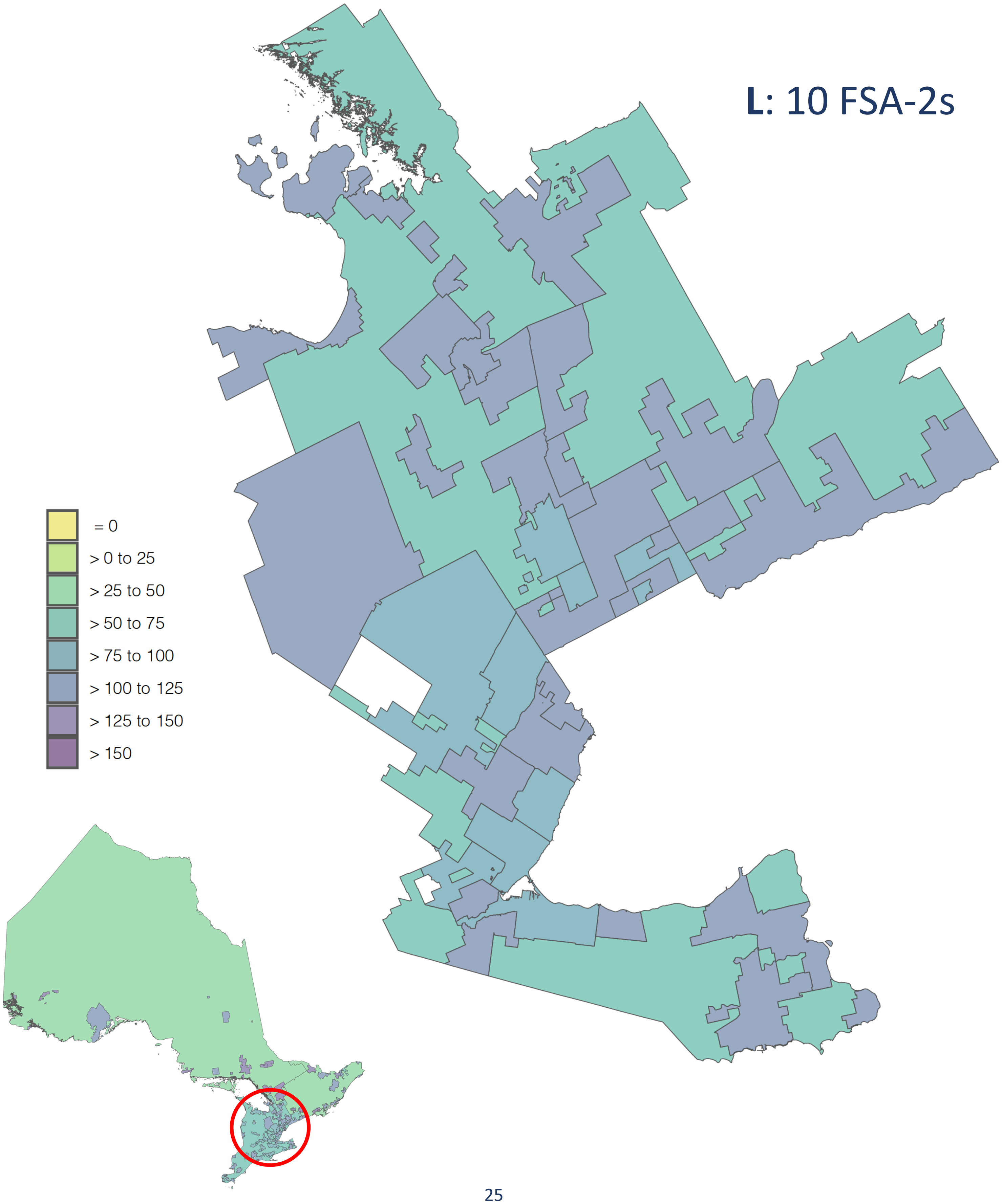

##### Heat Map 11: Dental Hygienists per 100,000 Population for M FSA-2s

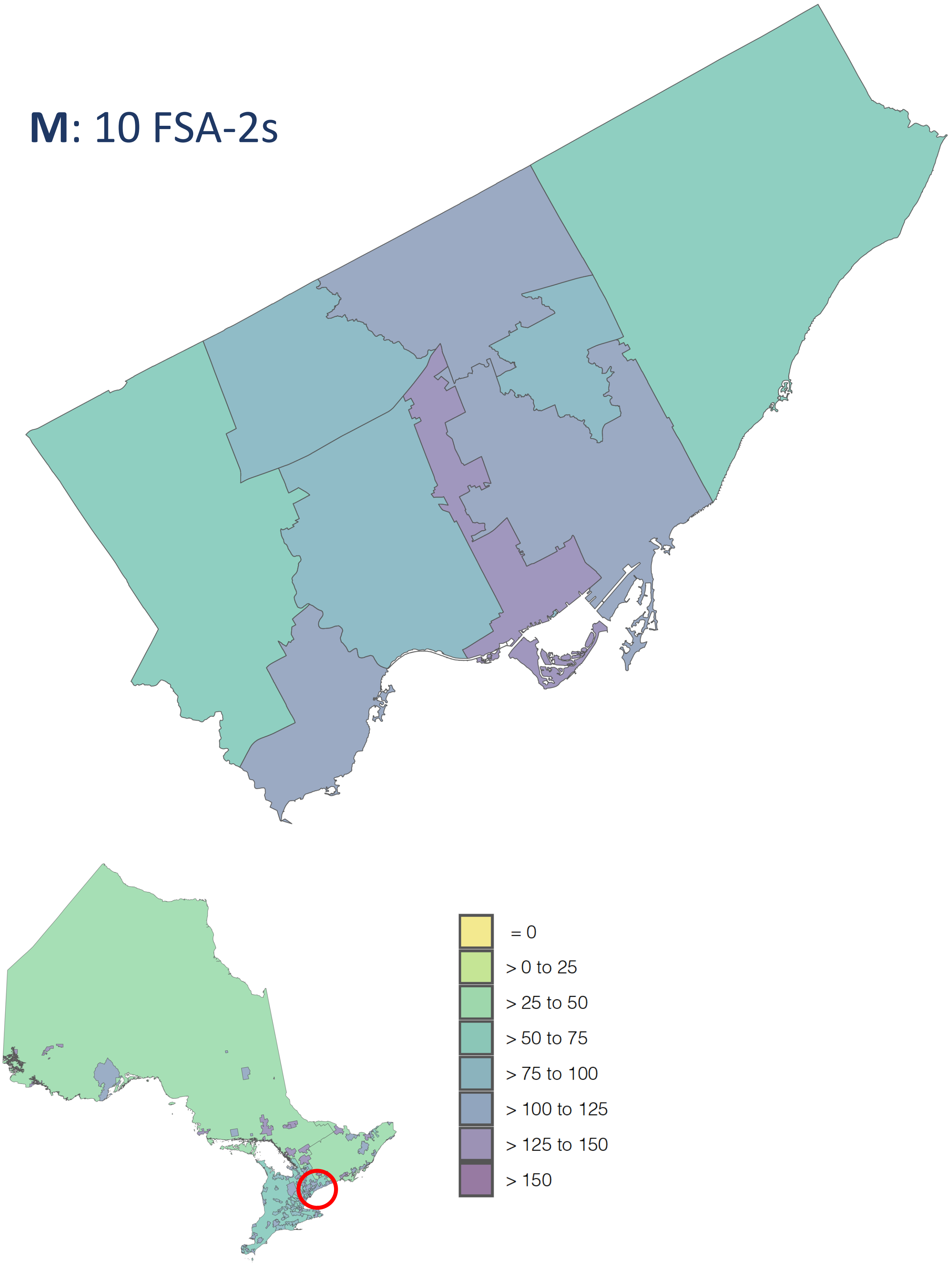

Heat Map 12: Dental Hygienists per 100,000 Population for N FSA-2s

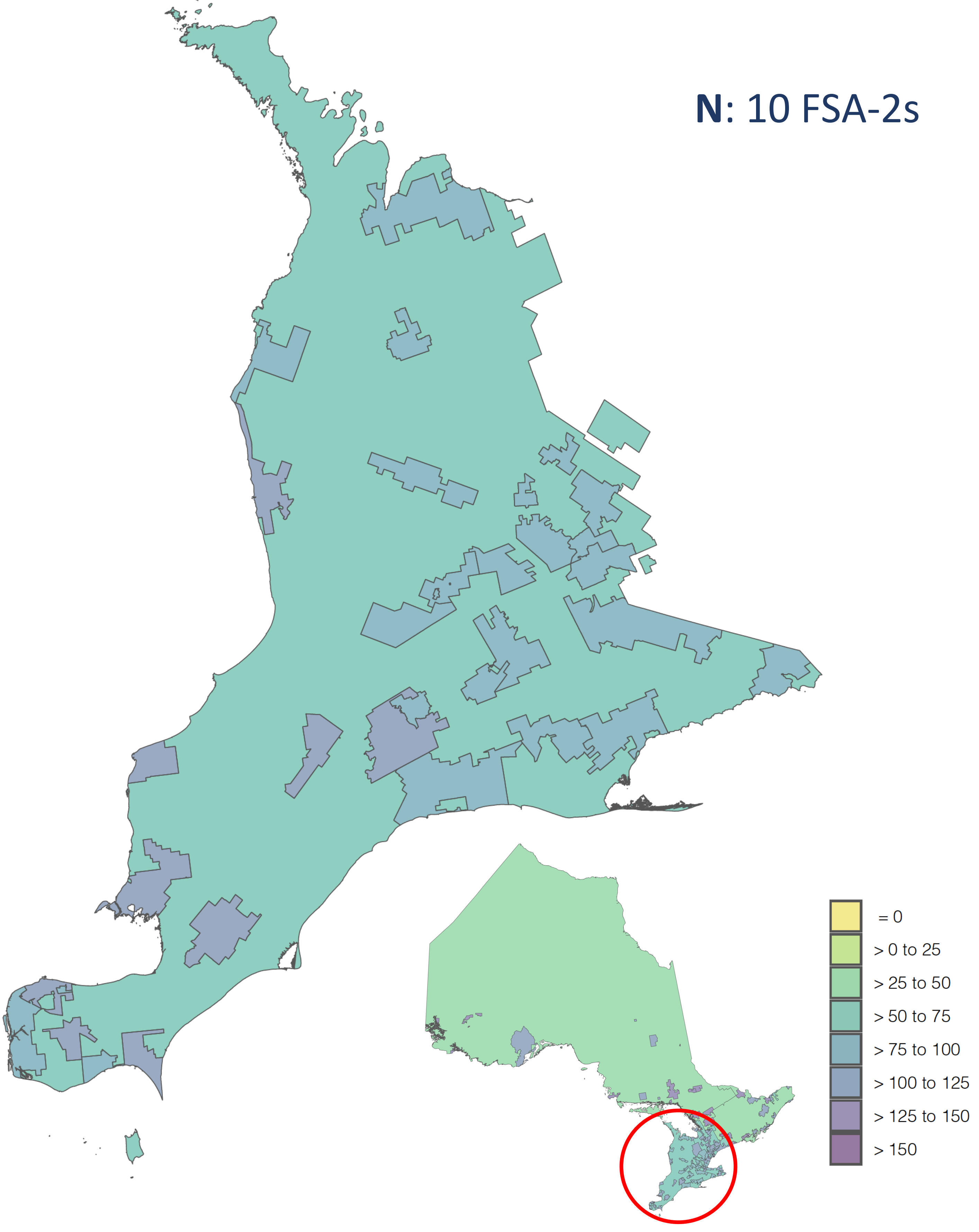

##### Heat Map 13: Dental Hygienists per 100,000 Population for P FSA-2s

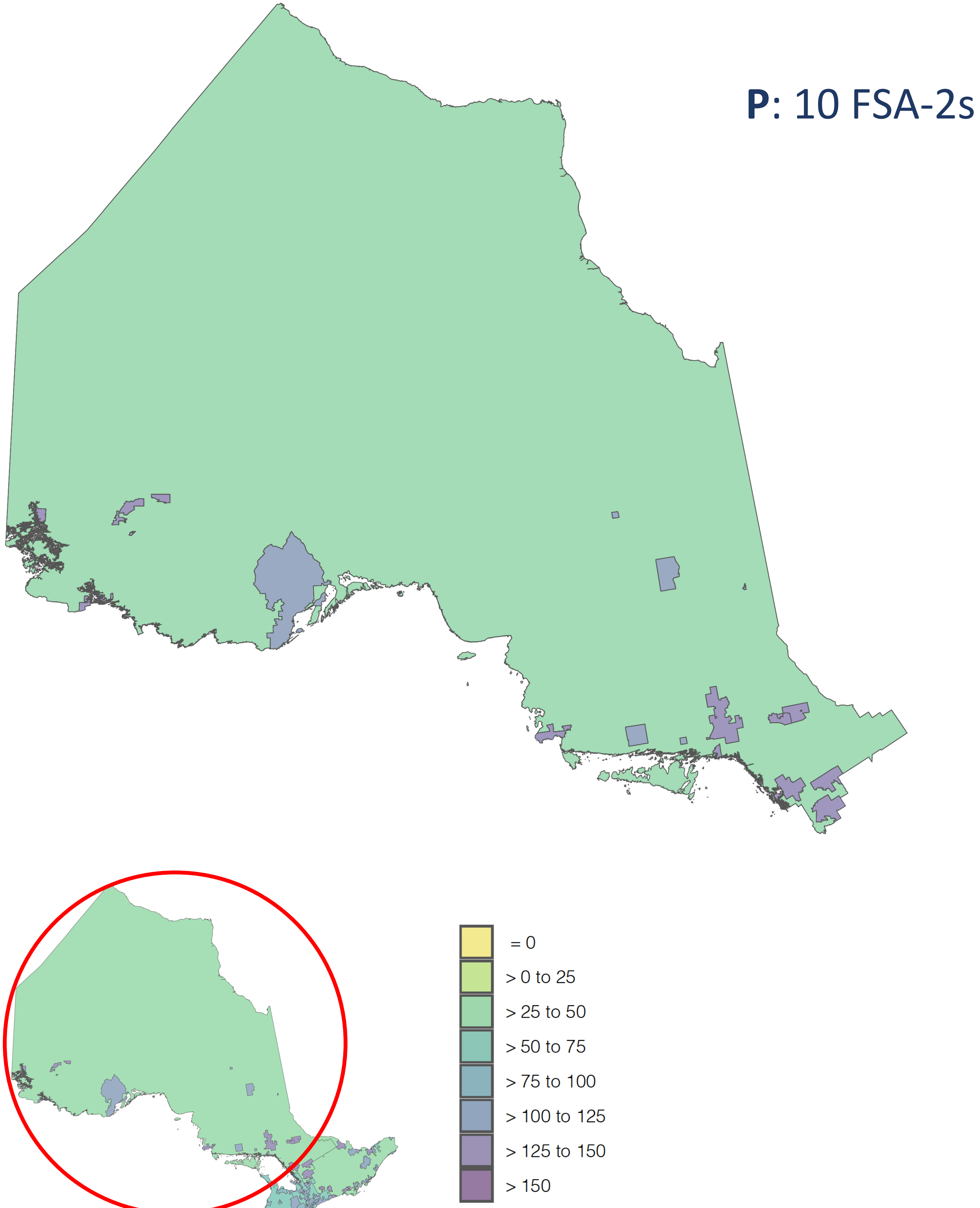
